# A semi-parametric, state-space compartmental model with time-dependent parameters for forecasting COVID-19 cases, hospitalizations, and deaths

**DOI:** 10.1101/2021.09.02.21262995

**Authors:** Eamon B. O’Dea, John M. Drake

## Abstract

Short-term forecasts of the dynamics of COVID-19 in the period up to its decline following mass vaccination was a task that received much attention but proved difficult to do with high accuracy. However, the availability of standardized forecasts and versioned data sets from this period allows for continued work in this area. Here we introduce the Gaussian Infection State Space with Time-dependence (GISST) forecasting model. We evaluate its performance in 1-4 week ahead forecasts of COVID-19 cases, hospital admissions, and deaths in the state of California made with official reports of COVID-19, Google’s mobility reports, and vaccination data available each week. Evaluation of these forecasts with a weighted interval score shows them to consistently outperform a naive baseline forecast and often score closer to or better than a high-performing ensemble forecaster. The GISST model also provides parameter estimates for a compartmental model of COVID-19 dynamics, includes a regression submodel for the transmission rate, and allows for parameters to vary over time according to a random walk. GISST provides a novel, balanced combination of computational efficiency, model interpretability, and applicability to large multivariate data sets that may prove useful in improving the accuracy of infectious disease forecasts.

## 1 Introduction

Questions about the future trajectory of the COVID-19 pandemic were central to the personal [1] and public policy [2] decisions of most of the world in 2020. In principle, predictive models can be key tools for decision support during infectious disease outbreaks, but their value rests largely on the trustworthiness of their ability to capture the key relationships in complex systems subject to changing rules and conditions [3]. One way to evaluate the trustworthiness of such a model is to measures its predictive performance over short-term time horizons [4].

One of the major initiatives to develop models with short-term forecasting value is the COVID-19 Forecast Hub [5], which collected forecasts of COVID-19 cases, hospitalizations, and deaths over the course of the pandemic in a standard format [6]. A standard format allows for standard evaluation of the accuracy of models among participating groups. Standard evaluations have proven useful in accelerating progress in other areas of machine learning [7], and projects such as the COVID-19 Forecasting Hub seek to facilitate comparable progress in infectious disease forecasting. Such projects are becoming more popular. The COVID-19 Forecast Hub evolved from earlier projects to collect standardized forecasts of seasonal influenza (e.g., [8]). Although the Hub collects forecasts for U.S. jurisdictions, forecasts are contributed from teams from all over the world, and the U.S. Hub has been matched by Hubs collecting forecasts for European nations (https://covid19forecasthub.eu/) and regions of Germany and Poland (https://kitmetricslab.github.io/forecasthub/forecast).

The COVID-19 Forecast Hub not only makes it relatively easy to compare the forecasts of different models made during the pandemic, but also creates a set of benchmark forecasting tasks which models developed later can use to demonstrate their predictive value. Of course, making good forecasts after the data are available is not as impressive as doing so beforehand, but this exercise still has value for developing models for the next pandemic, or resurgence of COVID-19. A model that can provide good prospective forecasts should also be able to provide good retrospective forecasts.

The non-stationary nature of COVID-19 pandemic [9] presents a challenge for forecasting by undermining the invariance assumptions that are the basis of most forecasting methods [3]. Castle *et al*. [9] draw a contrast between simple statistical models, which may be readily adapted to the most recent observations in a time series, and epidemiological models, which—although invaluable for providing insight into the past—are not as readily adapted to the most recent observations and thus more prone to systematic forecasting errors when the data are non-stationary. Here, we present a hybrid model, Gaussian Infection State Space with Time-dependence (GISST), which aims to combine the interpretability [10] of epidemiological models with the robustness to non-stationarity of statistical models by allowing key parameters of the epidemiological model to be fitted to the most recent observations. This model was developed for forecasting COVID-19 cases, hospitalizations, and deaths at the state level in the United States. The GISST model is interpretable in the sense that it incorporates our understanding of the high-level relationships among cases, hospitalizations, and deaths and produces outputs such as the effective reproduction number and estimates of the number of currently infected individuals. Further it includes a regression model for the effects human mobility and vaccination levels on transmission and susceptibility, and it has the potential to be used to identify new correlates of transmission from a large panel of candidate variables. Figure 1 provides a graphical summary of GISST. We describe the model in detail in Materials and Methods. In Results, we present parameter estimates and evaluations of the model’s 1-day ahead forecasts and 1-4 week ahead forecasts for the state of California. We find that the GISST model’s forecast performance approaches and sometimes exceeds that of the COVID-19 Forecast Hub Ensemble in 1-4 week ahead forecasts. It does particularly well for the hospital admissions indicator, which may be the indicator whose forecasts are of the greatest practical value. The GISST model does this while having some advantages in computational efficiency and interpretability compared with the COVID-19 Forecast Hub Ensemble model.

**Fig 1.**
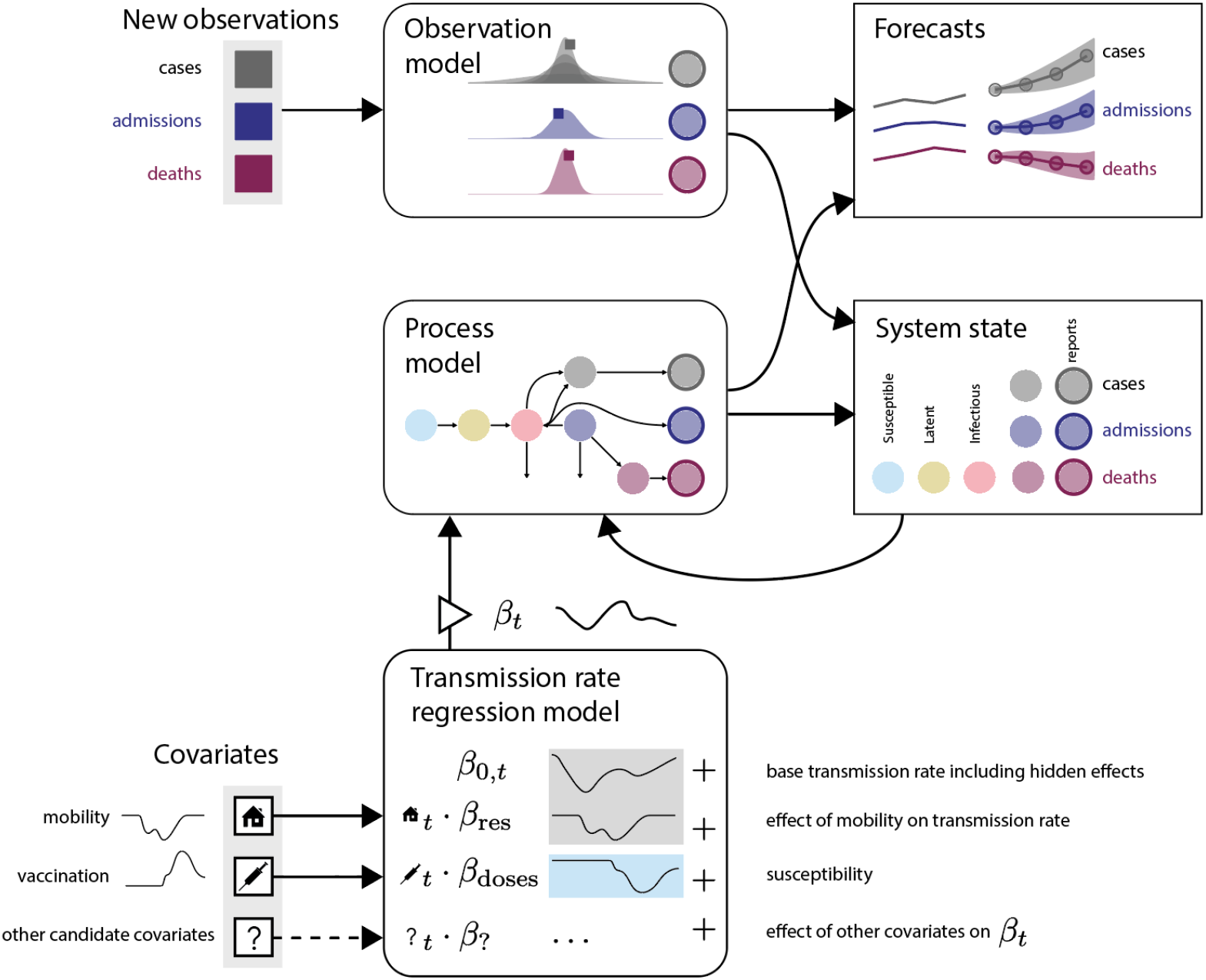
Graphical summary of the GISST concept for our modeling and forecasting infectious diseases. The main features are time-dependent parameters to accommodate the non-stationarity of COVID-19 dynamics, a regression model to incorporate data on covariates of transmission, and the integration of a compartmental epidemiological process model into a Gaussian state-space model for cases, hospital admissions, and deaths.

## 2 Methods

### 2.1 Data

#### 2.1.1 Forecasting targets

Many times series indicators of the status of the COVID-19 pandemic are available. To allow our model’s forecasting skill to be compared with those of many other models, we use the forecasting targets established by the COVID-19 Forecast Hub [5, 6]. These targets use data on cases and deaths distributed in the COVID-19 Data Repository https://github.com/CSSEGISandData/COVID-19 created by the Center for Systems Science and Engineering (CSSE) at Johns Hopkins University [11].

We also use data on hospital admissions in the “COVID-19 Reported Patient Impact and Hospital Capacity by State Timeseries” and “COVID-19 Reported Patient Impact and Hospital Capacity by State” datasets provided by the U.S. Department of Health & Human Services (HHS) on healthdata.gov. We accessed these data through the mirror accessible through the Delphi Epidata API [12]. Specifically, this time series is the sum of the fields labeled `previous day admission adult covid confirmed` and `previous day admission pediatric covid confirmed` in the tables provided by HHS.

One challenging feature of these time series in regards to forecasting is that they are subject to revision. To ensure a fair comparison between our model and those models that provided forecasts as the pandemic was progressing, for any forecast we used the version of the data available on the day for forecast submission to the forecasting hub. For evaluation of forecasts, we use a version of the data that is recent at the time of this writing.

The forecasting hub had a flexible submission format that allowed teams to submit for any subset of counties and states as well as the United States as a whole. We focus on targets at the state level in this work, in the largest state of California. One reason is that the county-level data are subject to more reporting anomalies. Another is that many important policy decisions are made at the state level. A third is that approximations used in our process model were better suited to modeling the dynamics of larger, state-level populations than those of smaller county populations.

The time series of cases, deaths, and hospitalizations are available at a daily resolution, but the targets are Sunday to Saturday weekly totals for cases and deaths. In keeping with the conventions of the COVID-19 Forecast Hub, we restrict our attention to 1-4 week ahead forecasts of cases and deaths and 1-28 day ahead forecasts of hospital admissions. The conventional wisdom is that forecasts at longer time horizons are generally not accurate enough to be useful [5]. Figure 2 shows the range of daily observation values of COVID-19 cases, hospital admissions, and deaths for California to illustrate the general trajectory of the time series used to fit our model as well as the extent to which it was revised over time.

**Fig 2.**
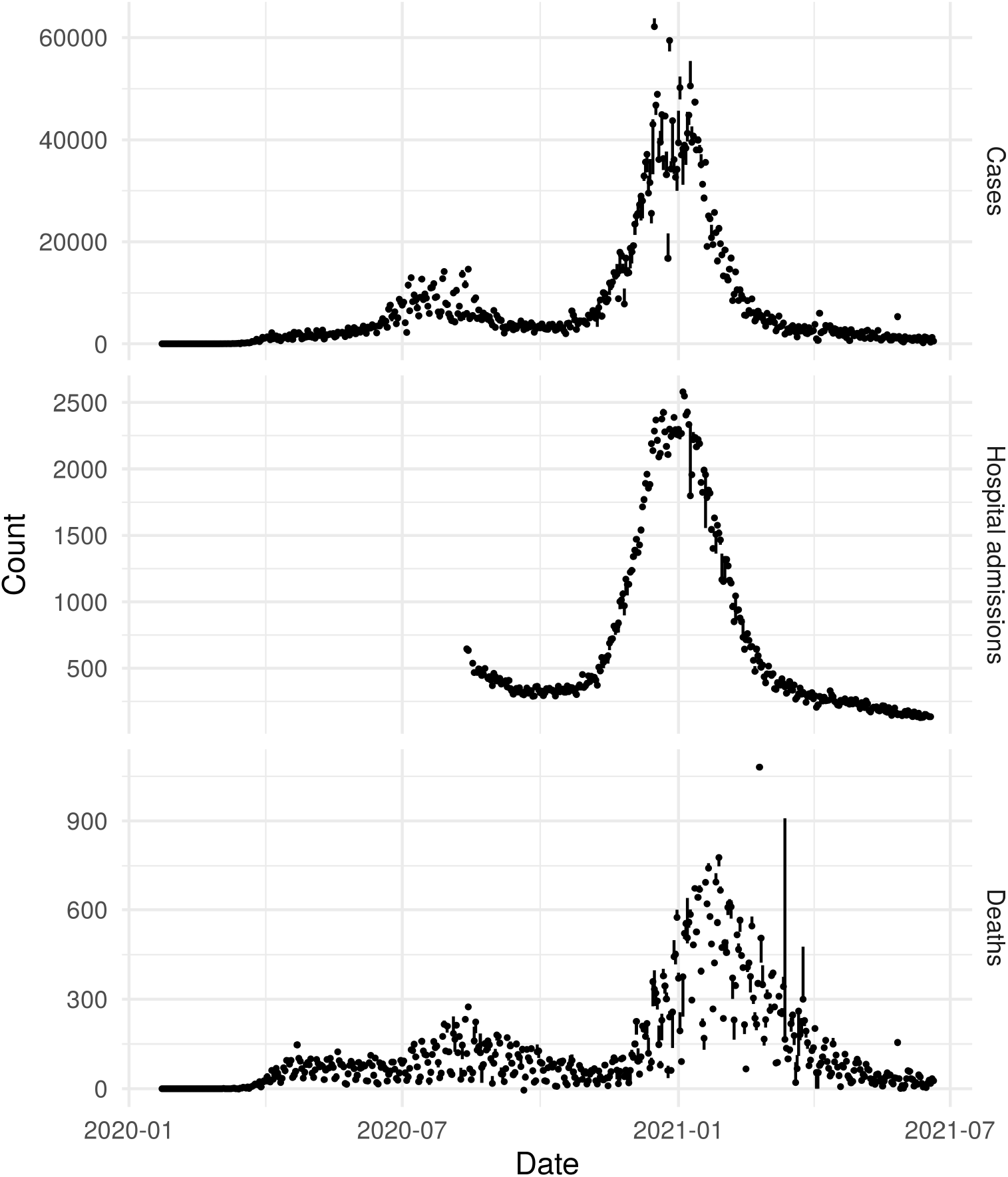
Time series of forecasting targets and range of revisions. Points are the California time series of COVID-19 indicators which our model is designed to forecast in the version of the data used to fit our model, which was the version available on June 21, 2021. The vertical line range gives the range of values that each observation had in all versions of the data used to fit our model, which were the versions of the data available on Mondays from June 29, 2020 to April 26, 2021.

The hospital admissions time series is unique in that it begins later than the cases and deaths time series, in the middle of summer, and in that it did not become available until November of 2020. At the beginning of the hospital admissions time series, there is also a misleading trend of growth that is simply due to the number of hospitals providing data increasing. To remove this artifact, we consider as missing the observations in which the field `previous day admission adult covid confirmed coverage` was below 90% of its last observed value. We apply this transformation both for the data sets used for fitting and those used for evaluation.

#### 2.1.2 Regression model covariates

The GISST model includes a regression which expresses the risk of infection to susceptibles in terms of likely predictors of immunity and exposure. One such predictor is an aggregated measurement of the amount of time individuals spend in residential areas, as quantified in Google’s Community Mobility Reports [13]. We use a rolling weekly average of the percent change from baseline in the original mobility reports. Figure 3 shows the resulting time series.

**Fig 3.**
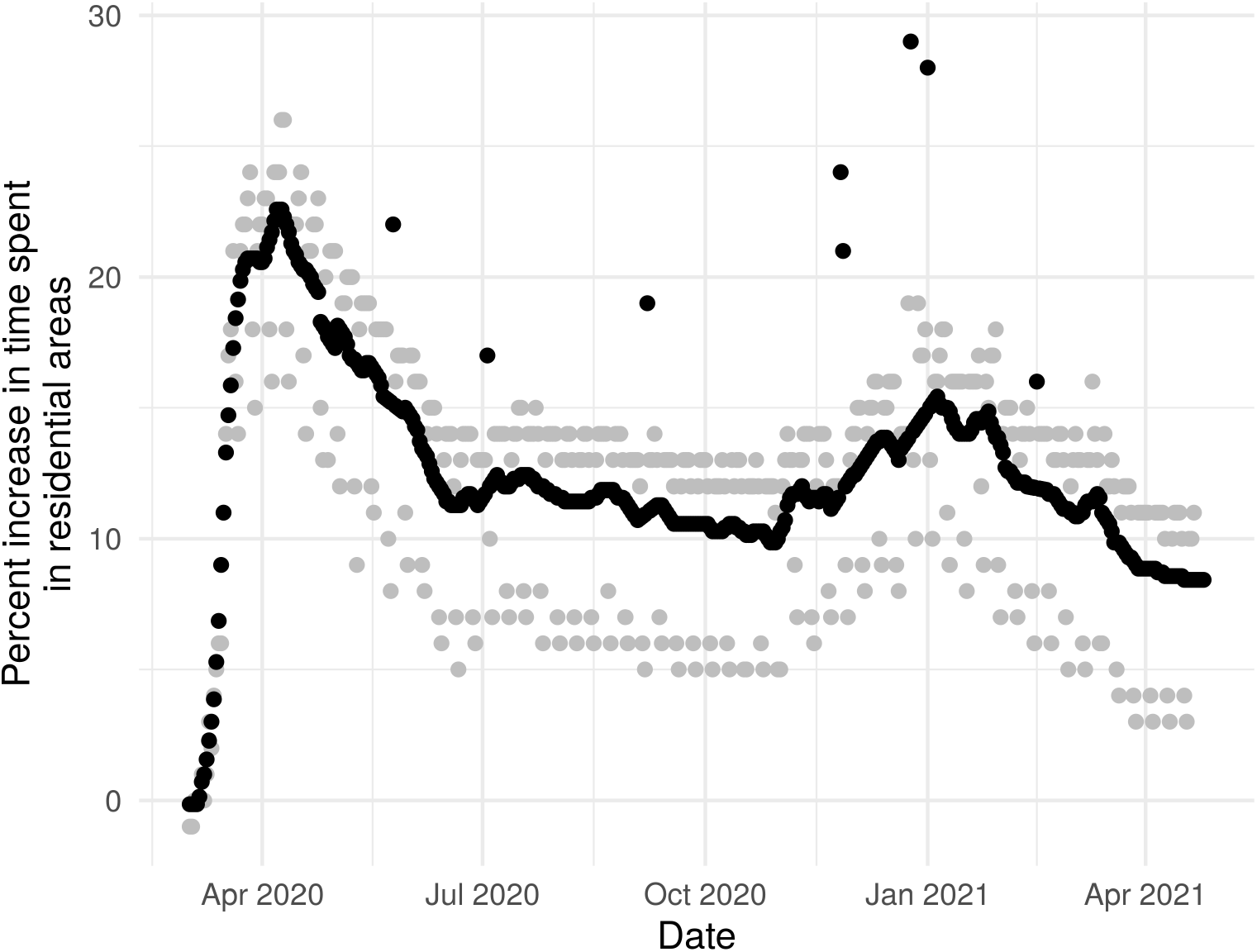
Indicators of the time spent at home. Grey points are the original data provided by Google of the amount of time people in California spent in residential areas relative to a pre-COVID-19 period. Black points are an example time series of the derived version which we use as a predictor of exposure in our model. Holiday effects in the original data are retained but weekly periodicity is removed. Consequently, the black and grey points coincide on the eight holidays considered in this time window. In other words, the value of this indicator on holidays is visible as the height of the black points separated from the main series of black points.

The details of the calculation of our mobility covariate are as follows. For a given forecast date, we obtained the latest snapshot of Google’s `Global Mobility Report.csv` file on http://web.archive.org that was made prior to the beginning forecast date in the UTC time zone. This was the the data available to all forecasting teams 22-23 hours before the submission deadline of 6PM ET. Although these data were not subject to revision, there was a gap of a few days between the last mobility data point and the forecast date which we are able to reproduce by using the archived data sets. To remove weekly periodicity, we calculated a moving 7-day average with a centered moving window. The last 3 observations of this 7-day average time series are missing because data for the full window is not yet available.

In model development, we noticed that holidays had a strong effect on the rolling weekly average. In particular, if the available data ended on a holiday, our practice of filling in the missing values at the end of the mobility time series with the last weekly average lead to inaccurate imputed values of this covariate. Because forecasts and parameters estimates could be quite sensitive to these imputed values, we developed the following method to deal more carefully with holidays. We considered as missing output from any windows which included the observed date of the holidays of President’s Day, Memorial Day, Independence Day, Labor Day, Thanksgiving Thursday and Friday, Christmas, and New Year’s Day. Missing values within the resulting 7-day average time series were imputed by linear approximation. The gap at the end of the time series was filled in by repeating the last non-missing value. Finally, values of the unsmoothed data on holidays were used to replace corresponding values in the smoothed time series. In this way, holidays do not affect the extrapolation of values in the gap between the end of the data and the forecast date, but they are still able to affect exposure in our model. Note, however, that we are only accounting for the reduction in exposure due to individuals being able to spend more time at home rather than any increased exposure due to social activity.

A second predictor of the risk of infection is the number of vaccine doses administered in a state. The modeling assumption is that average susceptibility declines with the number of doses administered. We model an effect on average susceptibility rather than number of susceptibles because a single dose of a COVID-19 vaccine does not generally fully immunize an individual. We sourced our time series of the number of doses administered from the Git repository at www.github.com/govex/COVID-19, which was created by Johns Hopkins University. These data came from either the states’ public dashboards or the CDC Vaccine Tracker. Since the reported numbers are cumulative, the creators of this data set used the larger of the values from those 2 data sources each day as the most up-to-date value. To obtain a covariate free of missing values, we assumed all values before the first reported value were zero and that all values after the last reported value were equal to the last reported value. Missing values within the time series were imputed with a linear approximation. Figure 4 shows the resulting time series.

**Fig 4.**
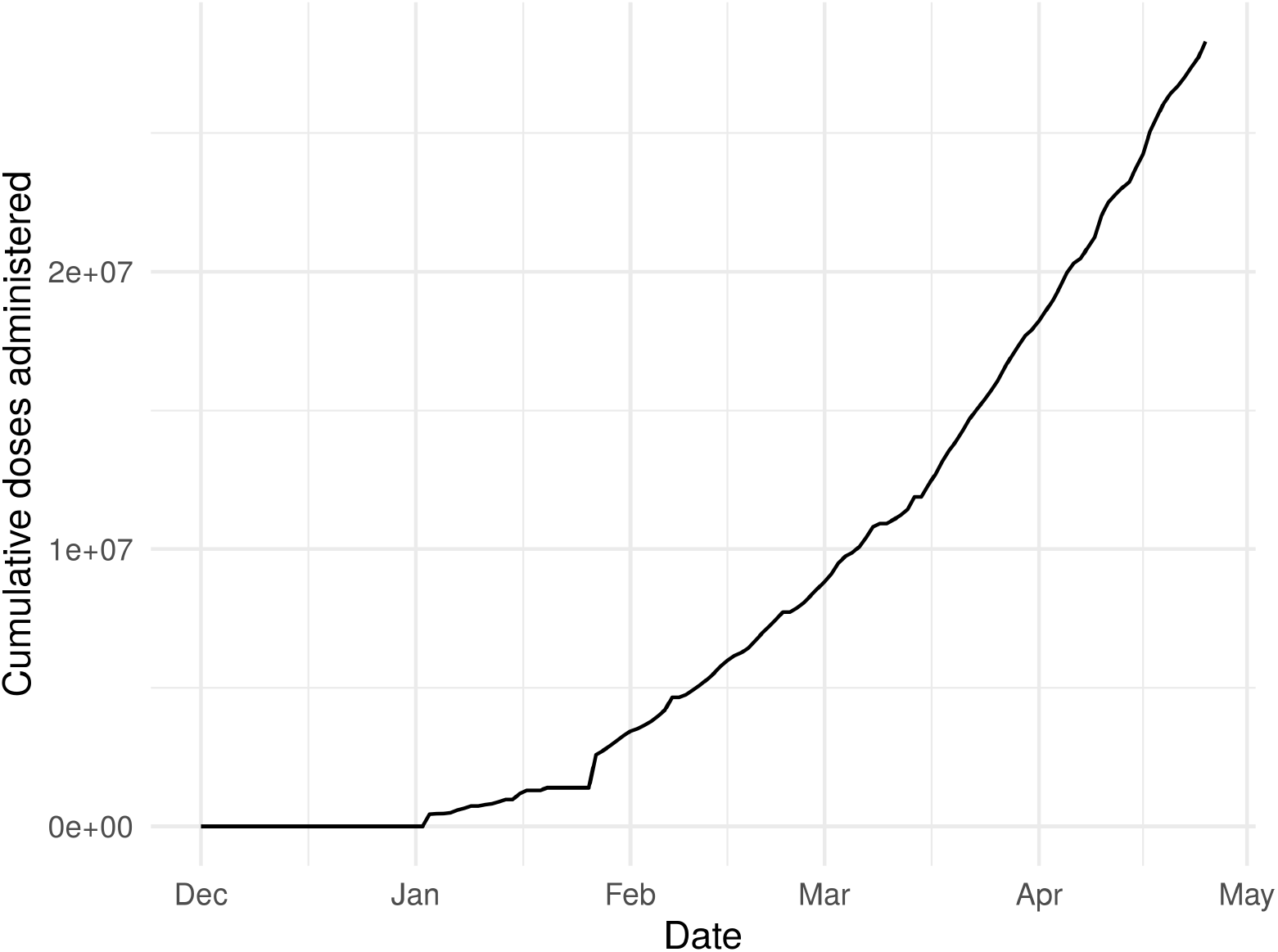
COVID-19 vaccination doses administered in California over time. This time series was used to estimate the effect of vaccination on reducing average susceptibility.

### 2.2 Model

The GISST model is a state space model, comprising a process model and an observation model. The process model and observation model both include stochasticity using Guassian random variables. For the process model, these Gaussian variables arise naturally by applying the system size expansion. We find the Gaussian assumptions justified by diagnostics of the fitted model, and it allows for the model’s likelihood to be efficiently calculated using the extended Kalman filter [14]. This efficiency makes it feasible to estimate the effects of covariates on the process model’s transmission rate in an embedded regression model, as well as allowing for parameters to vary over time according to a random walk to account for non-stationarity.

#### 2.2.1 Process model

Our process model arises as the large-population limit of a continuous-time Markov chain defined for the following state variables:

*X* Uninfected and *susceptible* individuals. Susceptible individuals can become infected by individuals in the *Y* compartment.

*L* Individuals with *latent* infections who are not yet infectious. At the end of the *L* stage, these individual progress to the *Y* stage.

*Z* Individuals who are *infectious*. This compartment includes both symptomatic and asymptomatic individuals. As a starting point, our model does not incorporate information about the extent of asymptomatic transmission and infectious individuals in our model may be thought of as a mean over all types of heterogeneity.

*Z*_*r*_ Individuals who have been diagnosed and will be reported as *cases*, but have not yet been reported. Those individuals are likely isolated and so are not infectious in our model. This compartment includes a count for the fraction of infectious individuals who recover without hospitalization, and it includes all infectious individuals who enter the hospitalized class.

*Z*_*r*_ This compartment keeps track of the number of cases reported each day, which we can compare with official reports.

*H* Individuals who have been *hospitalized*. Those individuals are modeled as not infectious due to isolation. A fraction of individuals in the *H* stage will recover, the remainder die of the infection.

*A* Individuals who are new hospital *admissions*. This compartment keeps track of the number of new hospitalized individuals each day, which we can compare with reported hospital admissions.

*D* Individuals who have *died* from the infection, but whose death has not yet been reported.

*D*_*r*_ The number of newly reported deaths each day.

The transition probabilities of the Markov chain are contained in Table 1. The definitions and values of model parameters appear in Table 2. The incubation rate used is consistent with an average incubation period of 4 days, which is a day shorter than the estimate of [15]. This shortening may be justified in light of the fact that that estimate is for the time of the appearance of symptoms, whereas we require the time until an individual becomes infectious and there is evidence that presymptomatic transmission is important for COVID-19 [16]. The removal rate *γ* was set to 4 based on the reasoning that it takes a few days for most individuals to obtain a test result once they become symptomatic and that infectiousness typically decreases over time following the appearance of symptoms [17], such that individuals are relatively unlikely to transmit after the first few days of their infectious period. This choice is consistent with the serial interval of 3 days estimated in ref. [16].

**Table 1.** Transition rules for the Markov chain.

| Event | Update | Propensity |
| --- | --- | --- |
| Transmission | $X \rightarrow L$ | $\beta_t XY/N$ |
| Incubation | $L \rightarrow Y$ | $\eta L$ |
| Removal of infectious individual whose cases will be reported | $Y \rightarrow Z$ | $(1 - p_{h,t})\rho_t\gamma Y$ |
| Removal of infectious individual whose cases will *not* be reported | $Y \rightarrow 0^*$ | $(1 - p_{h,t})(1 - \rho_t)\gamma Y$ |
| Reporting of cases | $Z \rightarrow Z_r$ | $\gamma_{z,w}Z$ |
| Hospitalization of an infection on a weekday | $Y \rightarrow H + A + Z$ | $p_{h,t}\gamma Y$ |
| Hospitalization of an infection on Saturday or Sunday | $Y \rightarrow H + A + Z$ | $p_{h,t}p_{h,\text{weekend}}\gamma Y$ |
| Death during hospitalization | $H \rightarrow D$ | $p_d\gamma_h H$ |
| Discharge from hospital | $H \rightarrow 0$ | $\gamma_h(1 - p_d)H$ |
| Reporting of deaths | $D \rightarrow D_r$ | $\gamma_{d,w}D$ |

\* The zero here indicates that we do not explicitly track individuals whose cases are not reported after they cease to be infectious. However, such individuals still contribute to the removed population which can be calculated as $N - X - L - Y$ .

**Table 2.** Parameters of Markov chain model.

| Symbol | Definition | Value |
| --- | --- | --- |
| $\beta_t$ | Transmission rate | Estimated |
| $N$ | Population size | Value given in JHU CSSE data set |
| $\eta$ | Incubation rate | 365.25 / 4 / year |
| $\gamma$ | Removal rate | 365.25 / 4 / year |
| $\gamma_{d,w}$ | Rate of reporting deaths on day of week $w$ | 365.25 / 10 / year * |
| $\gamma_{z,w}$ | Rate of reporting cases on day of week $w$ | 365.25 / 1 / year * |
| $\gamma_h$ | Rate of exit from hospital | 365.25 / 10 / year |
| $p_{h,t}$ | Probability of an infection leading to hospitalization | Estimated* |
| $p_{h,\text{weekend}}$ | Factor relating weekday hospital admissions to weekend admissions | Estimated* |
| $p_d$ | Probability of a hospitalization leading to death | Estimated |
| $\rho_t$ | Probability of a removal on day $t$ being reported as a case | 0.5* |

\* The parameter $p_{h,t}$ is fixed to 0.03 when hospitalization data are not available to avoid colinearity with $p_d$ . Additionally, $p_{h,\text{weekend}}$ is assumed equal to 0.9 when hospitalization data are not available. The parameter $\rho_t$ is estimated for dates up to June 2020 as described in *Estimation of $\rho_t$* , while $\gamma_{d,w}$ and $\gamma_{z,w}$ are fixed at this value for some days of the week and estimated for others as described in *Day of week effects on reporting*.

Our model diverges from the typical SEIRD (Susceptible-Exposed-Infected-Removed-Deceased) structure for specific reasons. A state variable for the number of removed individuals—that is, individuals who completed a pass through the infectious state—is missing because it would be redundant. That quantity can easily be calculated as *N – X - L -Y*, and the counts of deceased individuals, which are tracked by our model, subtracted from that if needed. The hospitalization reaction involves 4 state variables due to the nature of the data we observe. Hospitalizations are counted as both cases and hospital admissions. However, our observations of cases are reports which lag the diagnosis by several days whereas we have a time series of actual date of hospital admission. Further, some hospitalizations conclude in deaths, which will on average be reported with a longer lag from the date of hospital admission than will cases. Thus the potential to lead to observable events with three different lag times makes it necessary for the hospital admission event to propagate to three pathways in our model.

We could fit this Markov chain model using a simulation-based approach such as particle filtering. However, when the pandemic is in full strength and most state variables are large, it seems more advantageous to employ a central limit theorem to obtain a Gaussian distribution for the random fluctuations of the state variables. This choice allows for faster likelihood calculation via the Kalman filter. Therefore, computational resources may be redirected from calculating the minute details of our model’s stochastic fluctuations to other tasks important for developing good forecasts such as experimenting with variations of the model’s structure.

A Gaussian distribution for the state variables can be obtained by applying the system size expansion [18], which yields a system of ordinary differential equations for the expected values of the state variables and another system for the covariances of random fluctuations around the mean. The system size expansion has found use in earlier works in epidemiology for the analytic study of stochastic fluctuations of compartmental models [19, 20], but its use remains relatively rare in the field. As such, we provide some detail about about how the equations contained in the following paragraphs were obtained. In fact, the system size expansion provides many more results than we need. It provides equations for all terms in an expansion of the master equation for the Markov chain, but we require only use the two leading order terms, that is, the macroscopic equation and the linear noise approximation. So to facilitate exposition, we employ a less-used shortcut to these results by the simpler method of approximating the Markov chain in Table 1 with a chemical Lavengin equation [21].

The crux of the approach is to approximate the Poisson number of reactions occurring over a small time period with a Gaussian random variable. One advantage of using a Gaussian variable in place of a Poisson variable is that it allows for the techniques of stochastic calculus to be applied to characterize the system. The main advantage of the approximation for this work, however, is to allow the use of the Kalman updating equations to efficiently estimate our process model’s variables in consideration of both the the error in our observations and the stochastic nature of the process. The electronic supplementary material contains the equations used for these updates.

We next introduce the equations needed to calculate our model’s likelihood and offer remarks on their relation to the general derivation by Wallace [21]. As we have mentioned, we begin by writing a system of equations for the difference in each state variable over a small time step as a sum of Gaussian random variables. These variables represent the number of times each reaction in the system is fired over the course of the time step. The mean of this variable is set to the expected number of reactions as determined by the time step and the reaction rate in Table 1, and the variance is set equal to the mean to approximate a Poisson random variable. Next, the ansatz is made that the change in the variable may be decomposed additively into a deterministic solution which scales with the population size and a stochastic perturbation which scales with the square root of the population size. Then, the reaction rates are replaced with the Taylor series expansion of the reaction rates with respect to the stochastic perturbations. Collecting terms that scale with the population size leads to the following system of equations for the approximate expected values of the state variables:

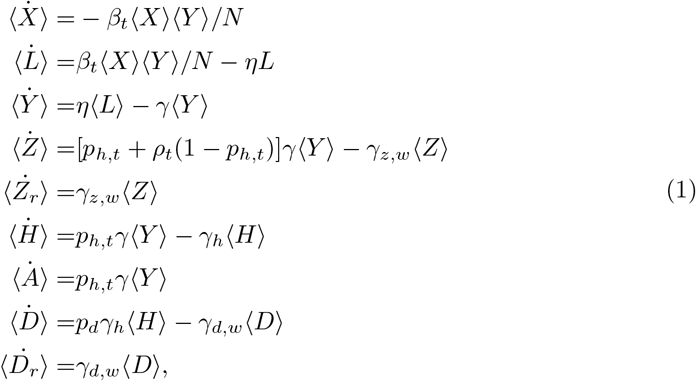

where the angular brackets indicate expected value and the overdots indicate time derivatives. Figure 5 provides a graphical representation of these equations. These equations are equivalent to equation 6 of ref. [21] multiplied by that reference’s system size parameter Ω. Collecting terms that scale with the square root of population size leads to the linear noise approximation for the stochastic perturbations. The equation for this approximation corresponds to equation 10 of ref. [21] multiplied by 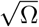.

**Fig 5.**
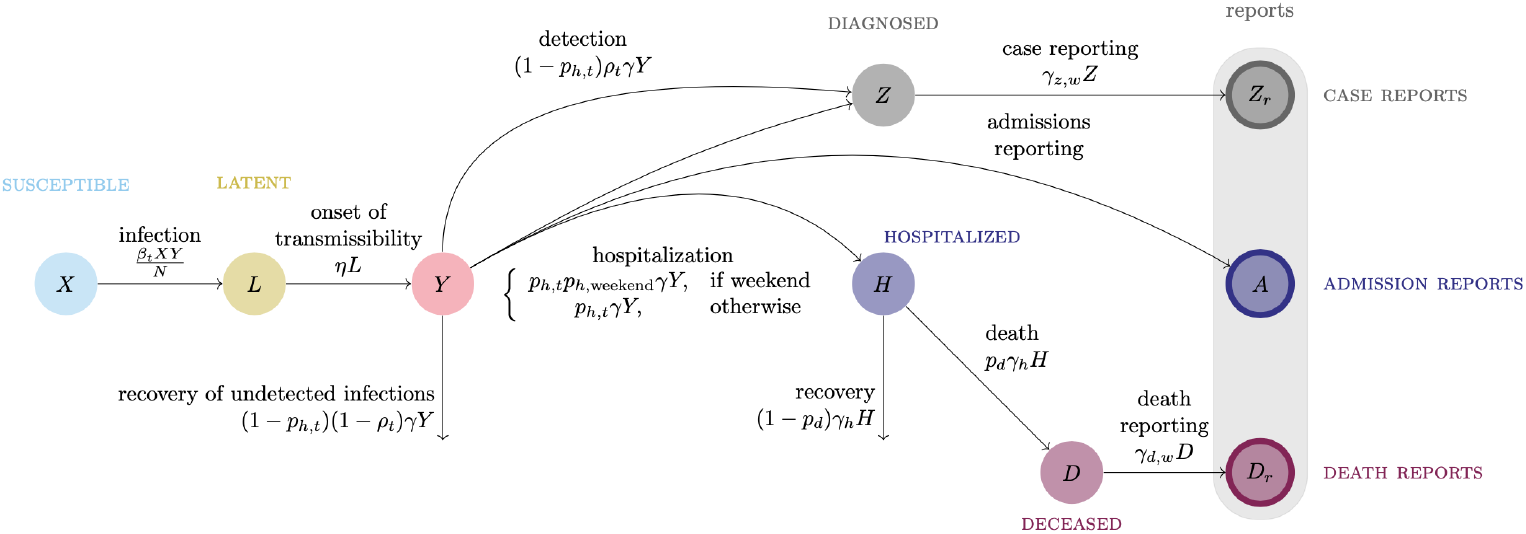
Flow diagram of differential equations for expected values of state variables.

Application of Itô’s isometry to that equation then yields a solution for the variance that is equivalent to equation 6.10 of Van Kampen [18]. Van Kampen [18] obtains this equation from his equation 6.9 via matrix algebra, the steps of which which may be reversed to obtain equation 6.9 from 6.10. Equation 6.9 in [18] corresponds to the next important equation for this work, a differential equation for the covariance matrix **P** of the stochastic fluctuations of the state variables::

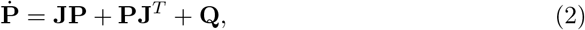

where **J** is the Jacobian matrix of the system in equation 1 and **Q** is the matrix defined by

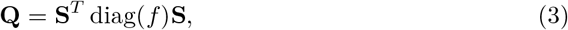

which may be calculated from individual-level reactions in Table 1. The matrix **S** has elements *S*_*ij*_, which are the stochiometric coefficients of species *j* in reaction *i* in the reactions in Table 1. The vector *f* collects the propensities column of Table 1. The solutions of equations 1 and 2 are the parameters of a Gaussian approximation to the distribution of the state variables in the original Markov chain. Thus, given an initial value of the means and **P**, numerical integration of equations 1 and 2 allows values of the mean and variances at a future time to be calculated. We describe how such values are used to generate forecasts and fit the model in the subsection *Fitting methods*.

We next explain some additional features of our model that proved necessary to obtain a good fit.

#### 2.2.2 Day of week effects on reporting

Although we do not attempt to model variation in exposure due to the day of the week, the effect of the day of the week on reporting of cases and deaths cannot be ignored. We account for this by allowing the reporting rate of cases, *γ*_*z,w*_, and the reporting rate of deaths, *γ*_*d,w*_, to depend on the day of the week. Rather than introduce a parameter for each day of the week, we break the days of the week into 2 or 3 groups and keep parameter values the same within a group. For *γ*_*z,w*_, there are two groups: (1) Saturday and Sunday and (2) Monday through Friday. For *γ*_*d,w*_ there are three groups: (1) Sunday and Monday, (2) Tuesday and Wednesday, and (3) Thursday through Saturday. These groups were chosen to parsimoniously account for day of the week effects by inspecting 1 day ahead prediction errors on different days of the week. The parameter *γ*_*z,w*_ was fixed to the value given in Table 2 on Monday through Friday while the weekend value was estimated. The parameter *γ*_*d,w*_ was fixed to the value given in Table 2 on Thursday through Saturday and its values on the other two groups of days were estimated. The parameter values in Table 2 were chosen to be broadly consistent with typical lags between case and death time series [22] and our analysis of individual-level data from the Georgia Department of Public Health.

Hospital admissions were also affected by the day of the week, being noticeably lower on weekends most of the time. We model this depression by introducing the parameter *p*_*h*,weekend_ with an allowed range of 0 to 1. As show in Table 1, the rate of hospital admission on weekends is calculated by multiplying the weekday rate by this reduction factor.

#### 2.2.3 Regression model for the transmission rate

Non-pharmaceutical interventions (NPIs) and vaccination have been effective in reducing the growth rate of COVID-19 cases, and we employ a regression model to allow for such effects in our model. Our covariates for quantifying these effects have been described in the *Data* subsection of *Materials and Methods*. We use doses_*t*_ to denote the number of vaccine doses administered on day *t* and residential_*t*_ to denote our indicator of the time spent in residential areas on day *t*. Our regression model for the transmission rate on day *t, β*_*t*_, follows

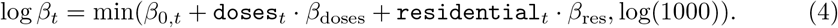

The upper limit of 1000 on the value of *β*_*t*_ reflects our scientific understanding that the reproduction number is unlikely to exceed 10 (which corresponds to *β*_*t*_ = 913) even in the absence of control measures and it prevents numerical problems in accurately calculating a solution for our process model with very large transmission rates.

Although for the sake of brevity we refer to *β*_*t*_ as a transmission rate, it serves to summarize effects on both transmissibility and susceptibility. That is *β*_*t*_ = (transmission rate on day *t*) *×* (suseptibility on day *t*). The intended interpretation of our regression model in terms of this factorization of *β*_*t*_ is

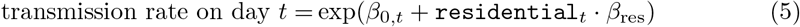

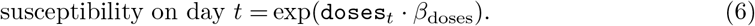

The coefficient *β*_0,*t*_ allows for the effects of NPIs for which we lack covariates to affect the transmission rate, and also allows for the reproduction number to decline with the depletion of susceptibles at a rate consistent with heterogeneity in susceptibilty [23]. These effects are allowed to vary over time according to a random walk model, which we describe in the subsection *Time dependence of parameters*.

#### 2.2.4 Observation model

Our process model accounts for variation around expected values of cases, hospital admissions, and deaths due to the intrinsic randomness of disease transmission. Another major source of variation comes from our imperfect observation of these variables. For the sake of simplicity and computational efficiency, we assume that these errors are zero-mean Gaussian random variables and are independent of each other. The variance of the observation error of hospital admissions, denoted *τ*_*h*_, and that of deaths, denoted *τ*_*d*_, are assumed to be constant over time. In contrast, allowing for variation in the observation error of cases over time proved critical to obtaining a good fit. For this variable, we assumed that variance of the observation error on day *t* is proportional to the model’s estimate of the number of infected individuals at the beginning of day *t*. This allows for a mean-to-variance ratio similar to what would result from assuming each case was reported with a given probability. Additionally, we allow the proportionality constant, denoted *τ*_*c,t*_, to change over time according to a random walk model. This allowed observations to be given less weight in periods subject to large amounts of noise.

#### 2.2.5 Time-dependence of parameters

The curve of cases in figure 2, with its multiple peaks, looks nothing like the typical simulation of our process model with fixed parameters. Even after incorporating the covariates in our regression model for the transmission rate, we, like other modelers [24], found it necessary to allow the parameters of our model to change over time to obtain a good fit. Further, allowing variation in parameters is justifiable given the rapidly evolving epidemiology of COVID-19, in which human behavior has played a large role. We allowed some parameters in our model to vary over time by allowing the parameter values to vary according to a random walk. The use of random walks to non-parametrically model complex epidemiological time series has been successful in forecasting seasonal influenza at the state level [25]. Table 3 lists the parameters in our model which we allow to take random walks, along with the frequency of steps in the walks, and the standard deviation of the normally distributed step size. Some of these parameters are transformed for the purpose of allowing normally distributed step sizes while staying within their natural domains. The frequency of steps and the standard deviations were determined through trial and error to achieve a good fit to the data. That is, they were treated as hyperparameters which were considered fixed when optimizing the likelihood with respect to our regular model parameters.

**Table 3.**
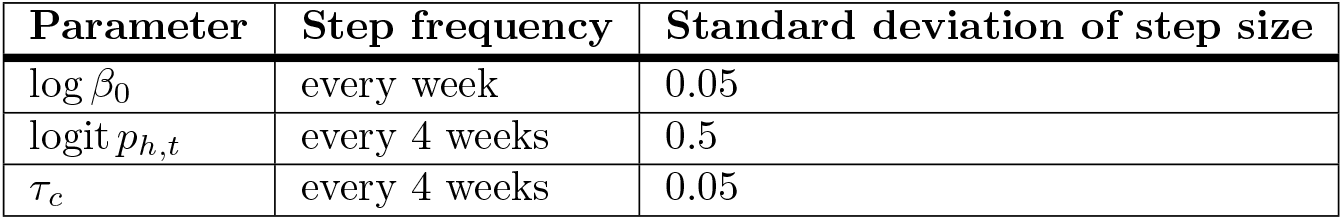
Random walk models for time-dependent parameters.

| Parameter | Step frequency | Standard deviation of step size |
| --- | --- | --- |
| $\log \beta_0$ | every week | 0.05 |
| $\text{logit } p_{h,t}$ | every 4 weeks | 0.5 |
| $\tau_c$ | every 4 weeks | 0.05 |

#### 2.2.6 Estimation of *ρ*_*t*_

Estimation of the time-dependence of *ρ*_*t*_, the reporting probability, required special treatment. This parameter changed the most in the first few months of the epidemic as testing volume increased. We accounted for this by estimating *ρ*_*t*_ for days before June 22, 2020, from the ratio of cases to deaths. This time window corresponds to the first major wave of cases. The idea is that most COVID-19 deaths have been reported with a relatively high and constant probability. We assume that the ratio of the probability of cases being reported to deaths being reported is increasing up to June 21, 2020, and plateaus at 0.5 on that date. This ratio is estimated by fitting a multivariate adaptive regression splines (MARS) model with the R package ‘earth’ [26]. The model is fit to the log of the ratio of cases to deaths 20 days later. We assume a monotonic increase of *ρ*_*t*_, and we ensure it by filtering the exponentiated prediction coming out of the MARS estimate through the cummax R [28] function. We further scale it so that the final value is 0.5, which is close to the estimate of the under-reporting factor for California in the same time period in the model of Irons & Raferty [27]. Figure 6 shows the resulting estimate of *ρ*_*t*_ for California.

**Fig 6.**
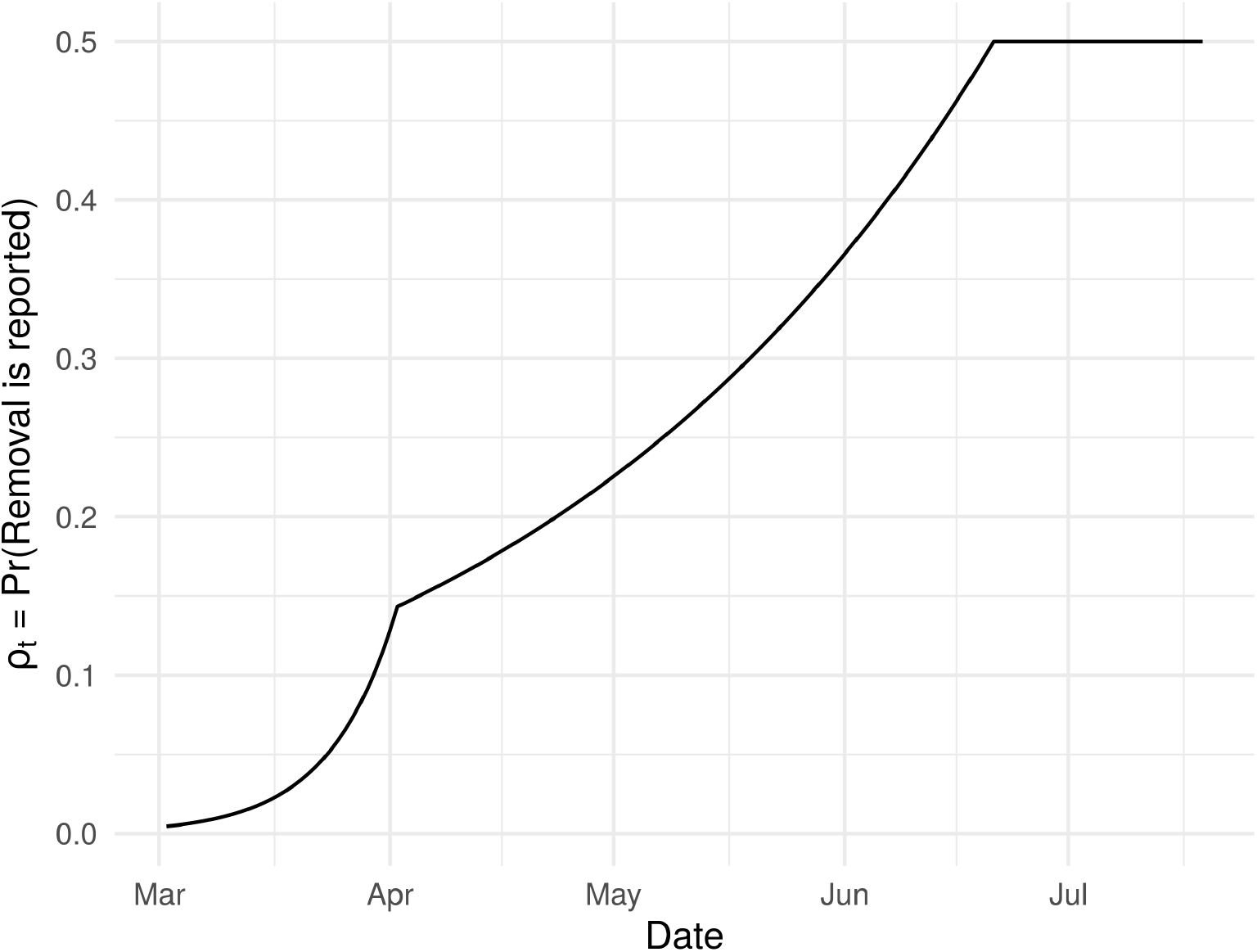
Estimate of the case reporting parameter *ρ*_*t*_ in California. This parameter is assumed to stay at 0.5 for dates beyond the last date plotted. See Tables 1 and 2 for the precise role of this parameter in our model.

### 2.3 Fitting methods

The GISST model has many parameters which must be estimated from the data before forecasts can be made. We do this using the method of maximum likelihood. The log likelihood of the model was calculated using an extended Kalman filter. The equations used in our extended Kalman filter implementation are nearly the same as those of the original Kalman filter introduced in Campagnoli *et al*. [14], the only difference being that our process model predictions are not generated by a linear function. We go through our likelihood calculation in detail in the electronic supplementary material.

#### 2.3.1 Optimization

For optimization of the log likelihood, we use the L-BFGS algorithm [29], a local optimizer. Local optimizers perform best when initialized in the neighborhood of the global optimum, so the first step in optimizing the likelihood was to choose good starting parameters for our optimizer. Out method for doing this is described in the electronic supplementary material. Initial parameters were iteratively improved by optimizing with the L-BFGS algorithm in the R package ‘lbfgs’ [32] to minimize the negative log likelihood. This algorithm was chosen for its ability to scale to a large numbers of parameters. Rate parameters were kept positive by fitting a log transform of them, and logits of probabilities were fitted to ensure they were valid probabilities. To keep all parameters on similar scales, the covariate doses_*t*_ was divided by *N* and the covariate residential_*t*_ was divided by 100. We provided a gradient function to the optimizer by using forward mode automatic differentiation via the ForwardDiff.jl [33] and DiffEqSensitivity.jl [34] Julia packages. Although most of our forecasting pipeline is written using the R language due to its many strengths for statistical modeling, we used Julia for the log likelihood function to make use of its robust numerical integration and automatic differentiation abilities. We ran the optimizer for 1000 iterations when fitting the first data set and when fitting the hospitalization data for the first time and 100 iterations when using a warm start. This number of iterations was typically sufficient to reduce the norm of the gradient to be 10 times smaller than the norm of the parameter vector. We stopped early if the norm of the gradient was 100 times smaller than the norm of the parameter vector. The suitability of the returned parameters was evaluated by calculating the Hessian matrix with automatic differentiation and verifying that it was positive definite (*i*.*e*., the negative log likelihood was convex). The inverse of the Hessian matrix was also used as a covariance matrix to generate Wald-type interval estimates for parameters [35].

### 2.4 Forecasts

Once a model is fitted, generation of forecasts is straightforward. In fact, the 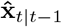 calculated as a part of the likelihood are already available as 1-step ahead point forecasts. We used these within-sample forecasts to evaluate the goodness of fit of a model. To generate out-of-sample forecasts, we simply simulated the process model in Eqs. 1 and 2 forward as far as needed beyond the day of the last observation and added the model-determined observation noise. In doing this, we used predicted values of *β*_*t*_ in the future from a lag-1 autoregressive (AR-1) model fit to the daily times series of the deviations of *β*_*t*_ from *γ* in the past. The model has its mean fixed at zero such that only the autoregression coefficient for the deviations is estimated. We denote this coefficient *ϕ* and provide an example of an estimate in Table 4. The input estimates of *β*_*t*_ have a similar time series to the estimates of *ℛ*_*e*_ plotted in figure 7. In this figure, one can see that following an initial transient period, *ℛ*_*e*_ had a tendency to fluctuate around and then revert to 1. This tendency may be a consequence of populations seeking to minimize the social and economic costs of non-pharmaceutical interventions (NPIs).

**Table 4.**
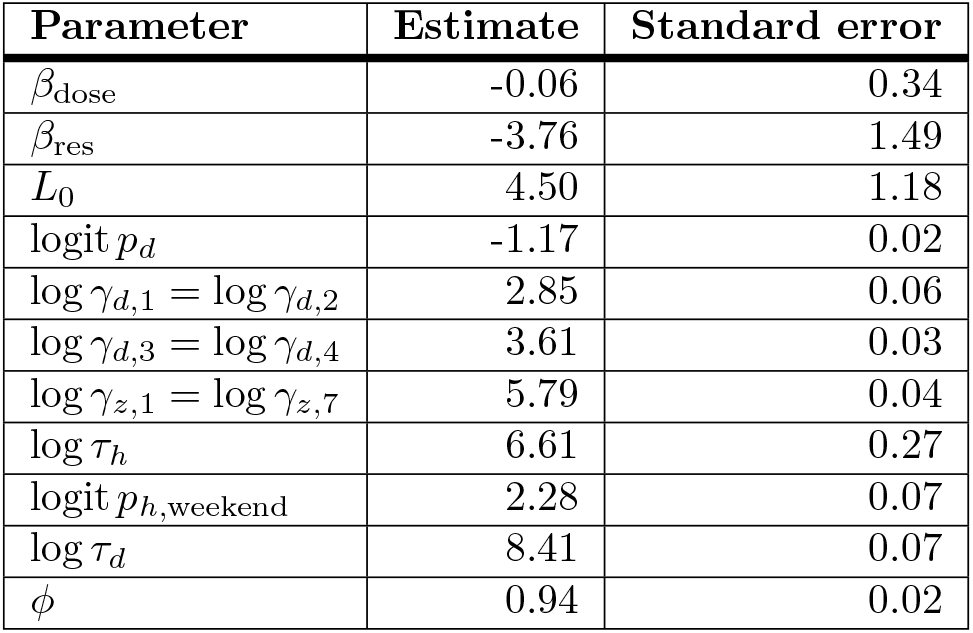
Maximum likelihood estimates of model parameters that are not time-dependent. Estimates are from the version of data available on April 26, 2021. *ϕ* is the coefficient for the AR-1 model of *β*_*t*_ used to make forecasts.

**Fig 7.**
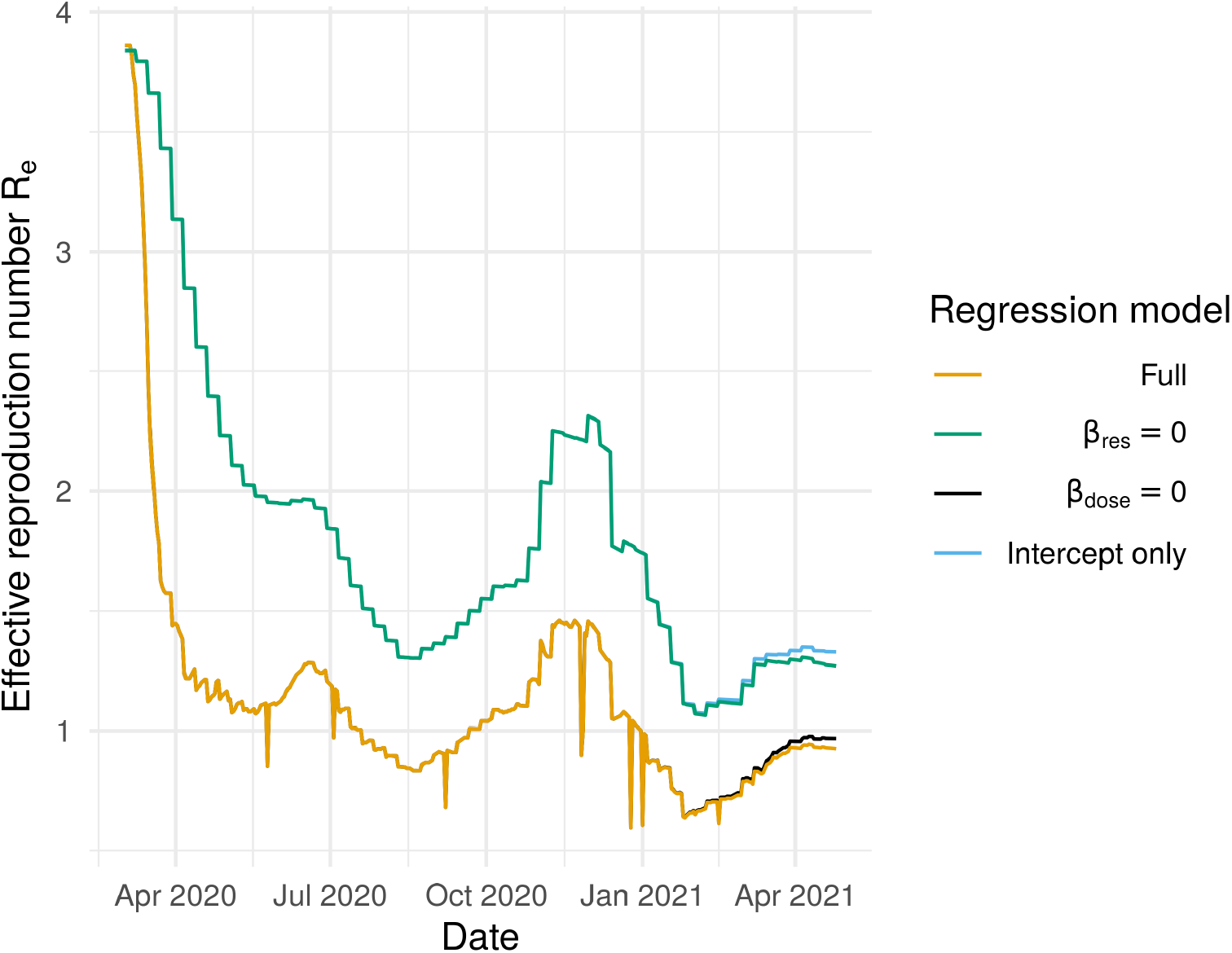
Effective reproduction number of fitted model for California. The model is fitted to data available on April 26, 2021. The full model corresponds to plugging *β*_*t*_ from equation 4 into *ℛ*_*e*_ = (*β*_*t*_*/γ*)(*X*(*t*)*/N*). We also plot *ℛ*_*e*_ with a *β* calculated by setting the effects of one or both of the covariates from their estimated values to zero to show their effects.

Perhaps NPIs are strengthened only when *ℛ*_*e*_ is clearly above one and COVID-19 indicators are on the rise and NPIs are relaxed soon after these indicators begin to drop. At any rate, we found that forecasts generated using predictions of *β*_*t*_ from an AR-1 model fitted to the estimates of *β*_*t*_ from days beyond May 1, 2020 up to the forecast date performed better than forecasts which assumed that *β*_*t*_ would remain at its most recently estimated value throughout the forecast period. The key benefit of the use of the AR-1 model seems to be that it limits the extent to which forecasts overshoot the peaks of pandemic waves.

In calculating forecasts, all parameters modeled as random walks were fixed at their final estimated value. To produce forecasts for the COVID-19 Forecast Hub, we recorded and zeroed the *D*_*r*_ and *Z*_*r*_ variables every 7 days to produce weekly forecasts of cases and deaths. Additionally, to calculate the observation model variance of a weekly observation, we summed the observation model variances that would have been applied to forecasts of individual days in the week. The Hub accepts probabilistic forecasts in the form of a set of 23 quantiles, {0.01, 0.025, 0.05, 0.1, 0.15, …, 0.85, 0.90, 0.95, 0.975, 0.99}, for deaths and hospital admissions and a set of 7 quantiles, {0.025,0.1, 0.25, 0.5, 0.75, 0.9, 0.975}, for cases. We obtained these quantiles using the qnorm function in R with the mean and variance of our model’s forecast distributions. Any negative quantiles—resulting from predicted distributions with a large coefficient of variation, for example—were replaced with zeros.

We conducted evaluations as follows. We used a version of data available on June 21, 2021. The extent of revision in our training data are summarized in figure 2. These revisions were large enough to decrease the accuracy of forecasts but not so large that the decreases were catastrophic. Forecasts were scored using the weighted interval score [36]. We scored forecasts of each indicator and forecast horizon separately. For each of these, we use a set of forecast dates for which our model, the COVID-19 Forecast Hub Ensemble model, and the COVID-19 Forecast Hub Baseline models have produced forecasts. The Ensemble model and Baseline models are useful reference points because the Baseline model represents a level of forecast skill achievable with little effort whereas the Ensemble model represents a model with a high level of forecast skill. The Ensemble model is formulated as the median value of the quantiles of forecasts submitted to the Forecast Hub and has performed better than most individual models. We analyzed forecasts of data from dates ranging from June 29, 2020, to April 26, 2021, which includes the second and third major waves of spread. We do not evaluate forecasts of the first wave of spread because Ensemble forecasts are not available for most of it. Additionally, the interpretation of the indicators in the first wave is not straightforward as many surveillance systems were changing rapidly as the severity of the pandemic grew. We do not evaluate forecasts beyond April 2021 because the indicator counts were reduced and subject to more changes in the reporting schedule as the pandemic began to wane.

### 2.5 Reproducibility

All data used in this study were obtained from public sources. An archive of the code used to obtain the data and generate all results is available on Zenodo at https://doi.org/10.5281/zenodo.5112578.

## 3 Results

### 3.1 Goodness of fit

We experimented with model structure until two goodness of fit criteria were satisfied. The first was that errors in the 1-step ahead forecast had the expected normal distribution. We used this diagnostic to evaluate whether that the model’s normality assumptions were reasonable. Figure S1 indicates that they were, with the exception of a relatively small number of outliers.

The second criteria was that MASE (mean absoluate scaled error) [37] was below 1 for all model fits. Due to the strong weekly periodicity in some of the data, we calculated MASE with both a non-seasonal naive model and a 7-day seasonal naive model. Our criterion was that both versions be less than 1. This condition provided a fast method to evaluate the forecasting skill of our model and its potential to outperform baseline models at the longer forecast horizons in our primary evaluation analysis. Figure S2 shows that our model for California satisfied these criteria.

### 3.2 Parameter estimates

An advantage of the mechanistic aspects of our model is that they provide some insight into why the a time series of indicators may be forecasted to go in a certain direction. To illustrate how this may be possible with our model, we next present an example of parameter estimates in our fitted models. The plausibility of these estimates in light of other sources of information about COVID-19 also provides a means to evaluate the assumptions of our model.

The effective reproduction number is perhaps the parameter of greatest interest in our model because it provides an easy-to-understand statement of whether the pandemic is growing or not [38]. The (instantaneous) effective reproduction number *ℛ*_*e*_ in our model is a derived quantity equal to *ℛ*_*e*_ = (*β*_*t*_*/γ*)(*X*(*t*)*/N*). Figure 7 plots the value of *ℛ*_*e*_ calculated from this expression with our model’s fixed values of *γ* and *N*, the predicted value of *β*_*t*_ resulting from our fitted regression model in equation 4, and the estimates of *X*(*t*) in **x**_*t*|*t*_ in equation S5 in the electronic supplementary material. The estimates seem reasonable, taking relatively large values early in the pandemic and then falling to and fluctuating around 1 after control measures were implemented. This figure also shows the contribution of individual regression model coefficients to the regression model’s prediction of *ℛ*_*e*_. All coefficients have effects in the expected direction, although the dose effect is small. Estimated values for these coefficients are in Table 4, along with estimates of other parameters that were not time-dependent.

Because we repeatedly fit the model to longer and longer time series for the purpose of making forecasts, even parameters which are assumed to be constant in a single model fit can reveal how the model fit or the underlying epidemiology changes over time. Figure S3 shows that the estimated effect of our indicator for the amount of time spent in residential areas was greatly reduced once hospital admissions data became available for fitting. We interpret this as a sign that our model was systematically incorrect in its assumed number of hospital admissions before being provided data, such that the availability of data triggered a step change in parameter estimates. Perhaps more epidemiologically interesting, the effect of this indicator trends toward zero from January onward. This behavior is consistent with NPIs becoming less important as vaccine-induced immunity built up.

Estimates of time-dependent parameters are shown in figures S4, S5, and S6. Overall, these parameters are estimated with a reasonable amount of precision and the trends seem reasonable. The probability of hospitalization is initially high and then quickly moves down to about 3 percent and fluctuates around that number. The rise in the random walk intercept *β*_0,*t*_ in the fall of 2020 coincides with the onset of the third major wave of cases. The observation variance in cases *τ*_*c,t*_ per infected individual grows in proportion to the number of cases reported. This variance-to-mean relationship reflects overdispersion relative to a Poisson model, which is frequently the case for observation models of infectious disease time series.

### 3.3 Evaluation of short-term forecasts

Weighted interval scores from our primary evaluation of forecast skill for cases and deaths are displayed in figure 8. The GISST model performs best relative to the reference models at the 1- and 4-week ahead horizon for cases and the 3-4 week ahead horizons for deaths. Due to the large scale of errors, the forecasts around the peak of the pandemic have the most weight in the overall score. It is thus possible to see how the overall scores in figure 8 are approximations of the average scores of observations in November through February in figure S7. In turn, the correspondence between these scores and the difference between forecasted and observed trajectories can be seen by comparing figure S7 with figures 9A and 9C. One can see how the good interval scores for the GISST model’s 4-week ahead deaths forecasts for observations in the final quarter of 2020 seen in figure S7 do indeed correspond to forecasts that are sharper and more accurate than the COVID-19 Forecast Hub Ensemble in figure 9C.

**Fig 8.**
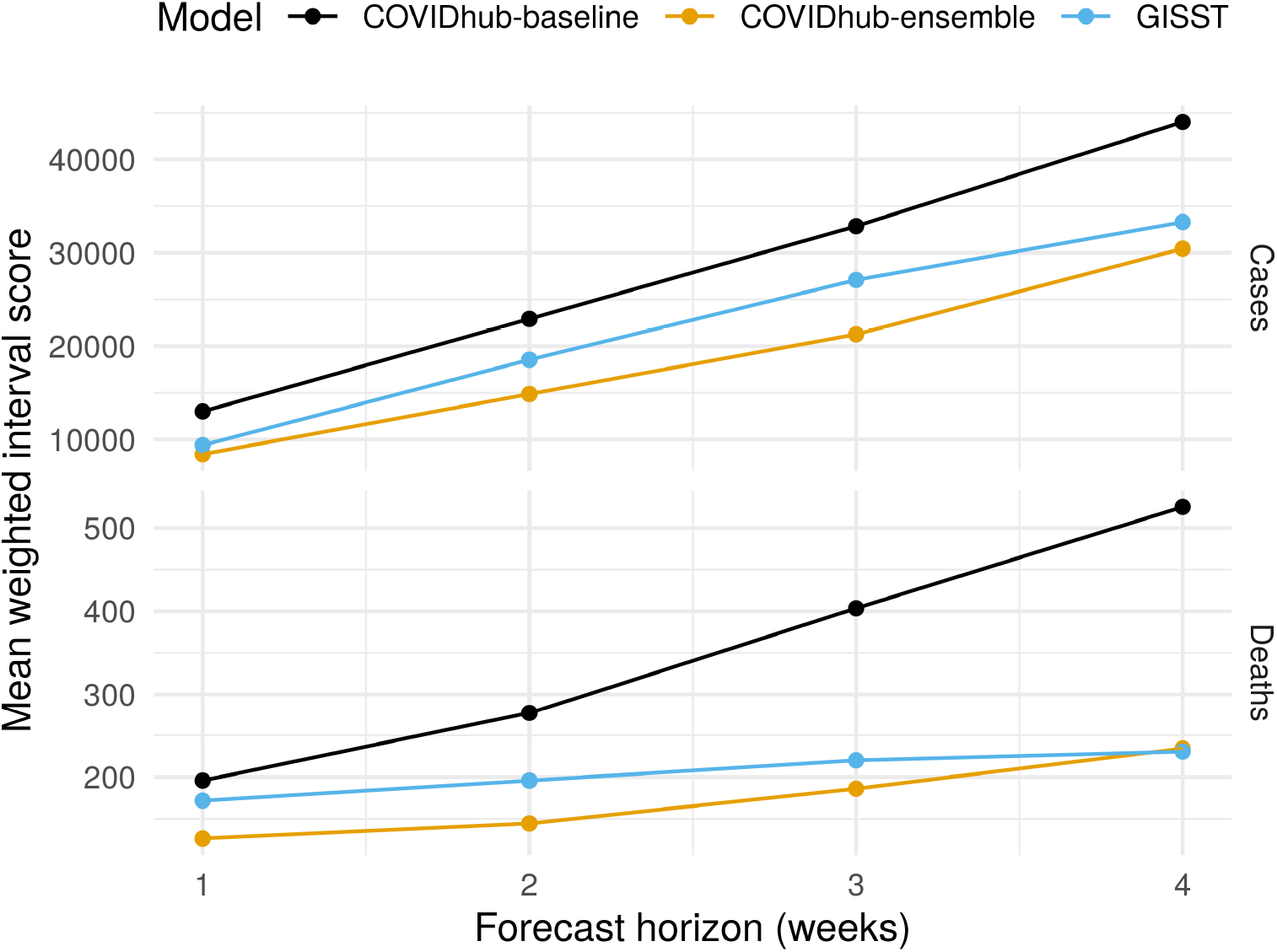
Overall performance of short-term forecasts of cases and deaths. Lower scores indicate better performance. The GISST forecast consistently outperforms the COVID-19 Forecast Hub Baseline forecast and performs relatively best for 4-week ahead death forecasts.

**Fig 9.**
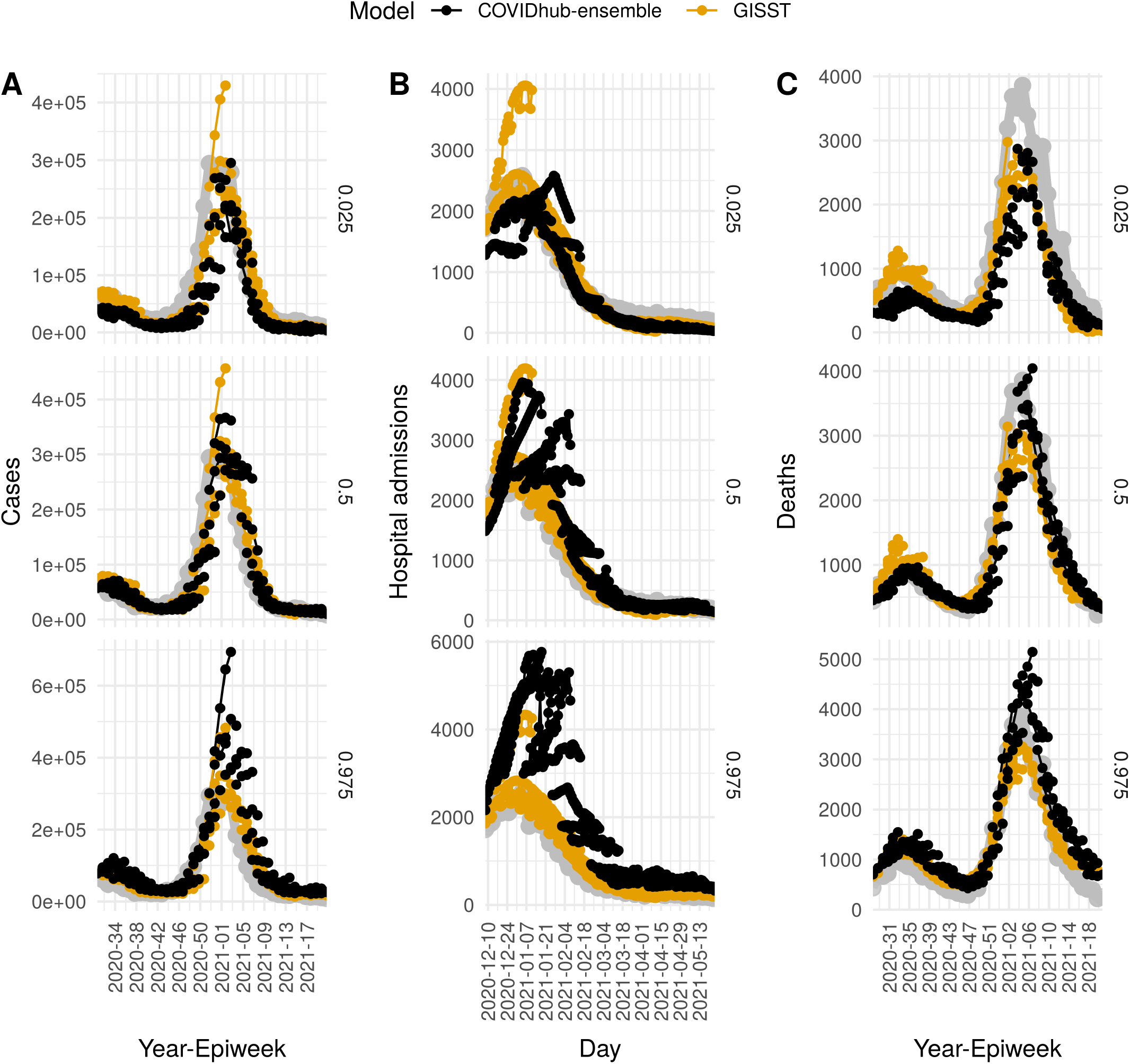
Forecast trajectories and target time series for cases (A), hospital admissions (B), and deaths (C). The panel labels at right are quantiles of the forecast. The target time series is plotted in grey. The performance penalty for underpredicting the true values is proportional to the quantile value, and the performance penalty for overprediction is proportional to 1 less the quantile value. Lines are drawn between forecasts of different horizons from a single forecast date.

Scores from our primary evaluation of forecast skill for hospital admissions are displayed in figure 10. The GISST model performs best relative to the COVID-19 Forecast Hub Ensemble model at all horizons except for 7 and 28 days. There is a clear periodicity in the weighted interval scores of the COVID-19 Forecast Hub Ensemble as a function of the horizon which likely originates from not modeling the reduction in hospital admissions on weekends (figure 10). Examination of forecast trajectories in figure 9B reveals that the overall better scores of GISST occurred in spite of poor forecasts early in the evaluation period, in which the 0.025 probability-level quantile of the the GISST forecast greatly exceeded the maximum value of hospital admissions in the December 14 forecasts. These poor forecasts were compensated for by consistent sharp and accurate forecasts from GISST for observations in the last half of January and throughout February. Performance at later dates was not as strong for GISST but these had less influence on the overall score due to the decreasing trend in hospitalizations.

**Fig 10.**
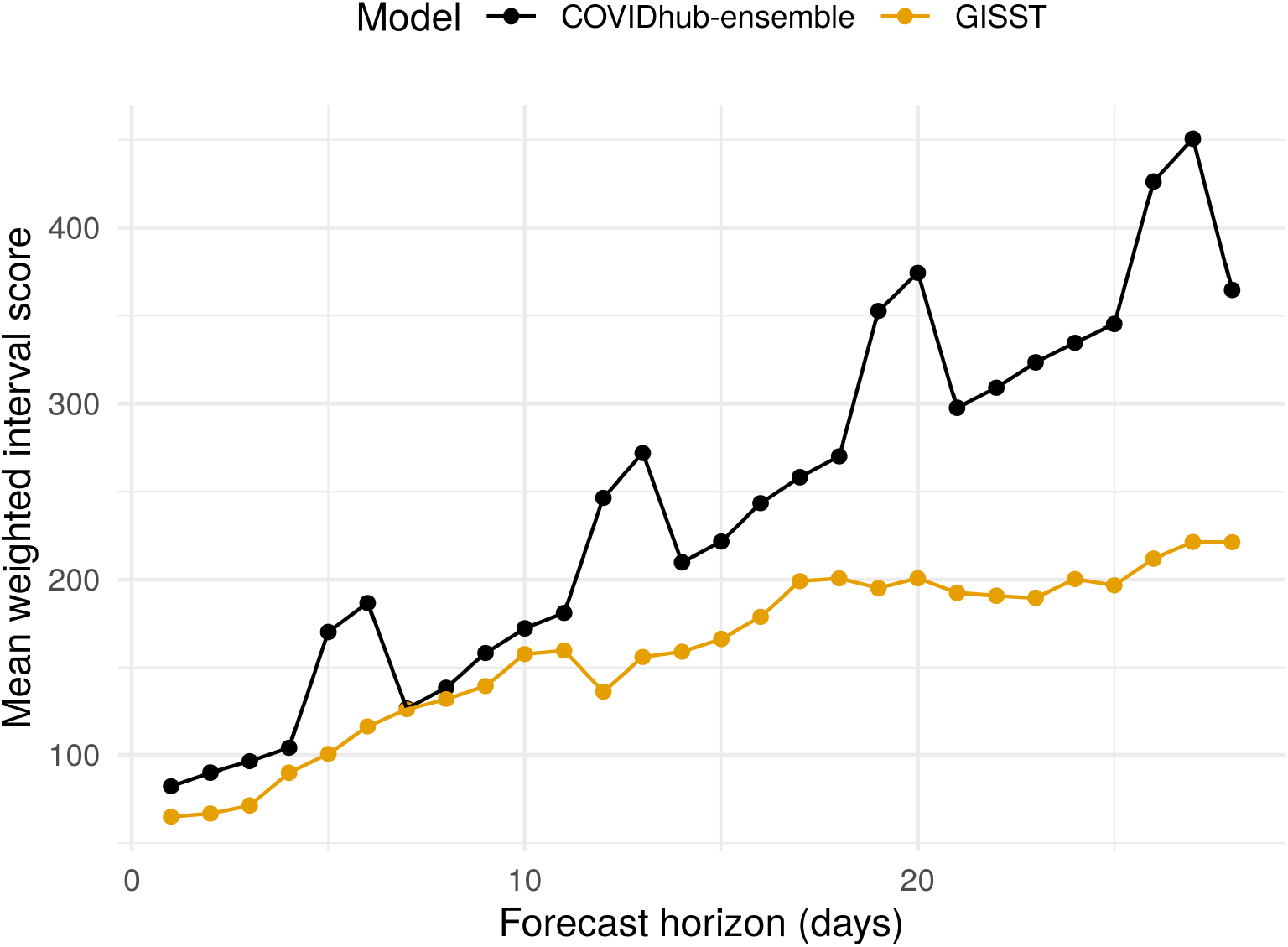
Overall performance of short-term forecasts of hospital admissions. Lower scores indicate better performance. The GISST forecasts score better than the COVIDhub Ensemble forecasts at all horizons. However, the difference in scores is slight at the 7 and 8 day horizons.

## 4 Discussion

We have introduced the GISST forecasting model and compared its performance in forecasting cases, hospital admissions and deaths with a simple baseline forecaster and a respected ensemble forecaster. Although some of its early December forecasts greatly overestimated future cases and hospital admissions, in summary evaluations of cases and deaths forecasts the GISST model dominated the baseline model and typically had a score closer to the ensemble model (figure 8). In summary evaluations of forecasts of hospital admissions, the GISST model scored better than the ensemble in 28 out of 28 horizons (figure 10). In addition to these positive performance attributes, GISST has the advantage of allowing trends in the forecasted indicators to be plausibly linked to each other and leading indicators such as metrics of human mobility. Additionally, it is computationally efficient. Even for the fits to April 2021 data sets, when our model had the largest number of parameters and observations, fitting an additional week of data required less than 4 hours of serial computation (figure S9) on a Linux system with an Intel Xeon E5-2450 Processor with a turbo frequency of 2.90 GHz. In addition to permitting forecasts to be generated online for a large number of locations, this efficiency facilitates model development by shortening the time required to test changes in model structure.

There are COVID-19 forecasts from many other forecasters besides the COVID-19 Forecast Hub Baseline and Ensemble models, and we next put our model in context with some of these other forecasters. The MechBayes model of Gibson *et al*. [24] provided accurate forecasts of COVID-19 deaths in real-time, and, like GISST, it does so by fitting a compartmental infectious disease model. But unlike GISST, the MechBayes model does not include a regression model for the transmission rate and requires Markov Chain Monte Carlo (MCMC) sampling to fit, which likely makes it more computationally costly. Further, it does not include hospital admissions. The method introduced in Srivastava *et al*. [39] is much more computationally efficient than GISST, but it does not forecast hospital admissions nor allow for transmission covariates. The Hub forecaster karlen-pypm [40] has produced good forecasts of all 3 indicators via a mechanistic population model. More recent versions of this model explicitly account for the dynamics of multiple variants of SARS-CoV-2. However, it assumes parameters are fixed except for at changepoints, which must be estimated.

Dehning *et al*. [41] present a fully Bayesian method for estimating such changepoints of transmission rates in a mechanistic population model. From the results of such estimation in time series of COVID cases in Germany, it seems that the data were only highly informative of about 1 out of 3 changepoint times. This result suggests to us that when prior information about the timing of changepoints is not readily available, a model that assumes a random walk in parameter values may be a better choice to avoid biases due to model misspecification.

Recently, Smith *et al*. [42] presented results from an SEIR-type model of COVID-19 in US states that included environmental covariates such as temperature and population density in addition to human mobility covariates. This was a spatially hierarchical Bayesian model fit with MCMC and likely more computationally demanding than GISST. Further, it forecasted deaths only and these forecasts were considered poor by the authors for horizons of more than 2 weeks. However, it is difficult to objectively compare accuracy with other models because the forecasts are not available in the standard Forecast Hub format.

The worst-scoring forecasts from GISST were made from dates in December (figures S7 and S8) when the GISST model greatly over-predicted the number of cases and hospitalizations (figures 9A and 9B). Although increasing the accuracy of such predictions may prove extremely difficult, future work should evaluate the benefits of incorporating uncertainty in future values of β_t_, which seems to be one of the strengths of the COVID-19 Forecast Hub Ensemble forecaster.

Another refinement to our approach which would likely lead to improved prediction of cases would be to allow for finer scale dynamics of disease transmission. For example, rather than modeling exclusively at the state level, a state-level model could be combined with models of groups of counties with similar dynamics. The county-level data of California show marked differences in dynamics relative to each other. For example, the relative height of the peak in cases in the 2020-2021 winter wave was much larger in the County of Los Angeles than it was in the City and County of San Francisco. Further, Bayesian fitting of a time-dependent SIR model to county-level data in South Carolina [43] revealed substantial heterogeneity in risk among counties as well as the value of a demographic feature (the percentage of the population below the poverty line) in explaining it. Modeling at the county level could thus allow for more accurate estimation of the number of susceptibles remaining in a population and their risk of infection. The state-space framework of GISST could readily accommodate the combination of models at different spatial levels via a hierarchical observation model.

Another limitation of the GISST model, as we have presented it here, may be that it is missing important covariates in the regression model for the transmission rate (equation 4). For example, we have not tried using other mobility metrics provided in the Google Community mobility reports. On the one hand, predicting future values of such covariates could be just as challenging as predicting future values of COVID-19 indicators without them. One might address this problem to some extent by applying a penalty proportional to the absolute value of the regression coefficient of these covariates, which would shrink the effect of unimportant covariates to zero. This approach would be readily achievable with the optimizer we used by choosing the Orthant-Wise Limited-memory Quasi-Newton (OWL-QN) algorithm instead of the LBFGS algorithm. The use of this option is what we alluded to in the introduction as the ability of GISST to search for important variables from a large space of candidate variables. Variables identified in this manner may sufficiently lead the indicators we wish to forecast that they improve forecasts just by being available up to or close to the date of forecast. Knowing which variables are important for predicting the transmission rate, even if they are difficult to forecast, could also be valuable for the design of forecasts.

## Supporting information

Electronic supplementary material

## Data Availability

All relevant data are publicly available at sources cited in the main text and all relevant
code is publicly available online.

https://doi.org/10.5281/zenodo.5112578

## Acknowledgments

We thank Éric Marty for creating the flow diagram of the ODE system and the graphical summary of our method.

