## Supplementary material for "A semi-parametric, state-space compartmental model with time-dependent parameters for forecasting COVID-19 cases, hospitalizations, and deaths": Electronic supplementary material

### 1 Log likelihood calculation

The first step in calculating the log likelihood is to initialize the means and covariance of our process model's state variables to their values on the beginning of the day of the first observation, which corresponds to March, 2, 2020. The initial value of $\langle L \rangle$ is a model parameter, which we denote $L_0$. The initial values of $\langle Y \rangle$, $\langle Z \rangle$, $\langle H \rangle$, and $\langle D \rangle$ are calculated from $L_0$ assuming that the time derivatives in equation 1 in the main text are zero, using the values of any time-dependent parameters when $t = 1$. This assumption is not intended to be highly accurate but rather to generate values that are plausible. The prediction-updates described later will serve to sharpen this initial estimate based on observations. The initial value of $\langle X \rangle$ is set to $N - \langle L \rangle - \langle Y \rangle - \langle H \rangle - \langle D \rangle$. The initial values of $\langle Z_r \rangle$, $\langle A \rangle$, and $\langle D_r \rangle$ are all set to zero, since these variables are meant to represent the accumulation reported cases, hospital admissions, and deaths over the coming day. Sometimes, during optimization, these initial values can become unreasonable and lead to numerical problems in calculating the likelihood. To avoid such problems, we do not allow any values to be greater than $N$, and we do not allow the intial value of $\langle X \rangle$ to be less than $N/10$. All of these initial values for means are collected in a vector denoted $\hat{\mathbf{x}}_{0|0}$. This is the initial vector of means for our process model. The initial covariance matrix for this model is denoted $\mathbf{P}_{0|0}$ and is set to a diagonal matrix in which all elements are 1 except for those corresponding to $\langle Z_r \rangle$, $\langle A \rangle$, and $\langle D_r \rangle$. These values are equal to zero at the beginning of the day by definition so their variances are set to zero. Similar to the means, the covariance matrix estimate is simply a rough approximation which the algorithm will sharpen. With the process model fully initialized, the next step is to generate a predictive distribution for the observed data based on the process model.

The predictive distribution for the next observations are generated by numerical integration of the system of equations in equations 1 and 2 over a time span of 1 day, where we used the convention of 365.25 days per year. For this step we used the `Tsit5` [1] integrator in the DifferentialEquations.jl package [2]. When solved with starting values from time $t$, the resulting vector of means is denoted $\hat{\mathbf{x}}_{t+1|t}$ and the resulting covariance matrix $\mathbf{P}_{t+1|t}$. If any elements of $\hat{\mathbf{x}}_{t+1|t}$ are negative, this must be due to numerical errors, so we set them to zero. Similarly, if any negative values appear on the diagonal of $\mathbf{P}_{t+1|t}$, the corresponding rows and columns are set to zero. The next step is to iteratively update these predictions for the mean and covariance based on the observed data.

The update of the prediction is done by using equations that minimize squared prediction error when the process model is linear, and which remain relatively accurate when the model is non-linear. An intermediate step in this update is to calculate the Kalman gain matrix $\mathbf{K}_t$. This matrix determines the weight to give to two types of predictions: (1) predictions of the state variables on day $t$ calculated as described in the previous paragraph by projecting past estimates of the state variables forward according to the process model, and (2) predictions of values of the state variables on day $t$ based solely on observations on day $t$ and the observation model. The equation for $\mathbf{K}_t$ is

$$\mathbf{K}_t = \mathbf{P}_{t|t-1}\mathbf{H}^T\mathbf{\Sigma}_t^{-1}, \tag{S1}$$

where $\mathbf{H}$ is a matrix that maps the state variables to observed variables and $\mathbf{\Sigma}_t$ is the sum of the covariance matrix $\mathbf{R}_t$ of the observation model and the projection of the covariance matrix of the process model into the observed coordinates. The matrix $\mathbf{H}$ is $3 \times 9$ and contains zeros except for one column in each row, which corresponds to the state variables $Z_r$, $A$, and $D_r$ respectively in rows 1, 2, and 3. The matrix $\mathbf{R}_t$ follows

$$\mathbf{R}_t = \begin{bmatrix} \tau_{c,t}\langle Y\rangle_{t-1} + 1 & 0 & 0 \\ 0 & \tau_h + 1 & 0 \\ 0 & 0 & \tau_d + 1 \end{bmatrix}, \tag{S2}$$

where $\langle Y\rangle_{t-1}$ is the estimate of $Y$ in $\mathbf{x}_{t-t|t-1}$. The ones on the diagonal of $\mathbf{R}$ prevent numerical problems that could occur if the $\tau$ parameters are all nearly zero. The matrix $\mathbf{\Sigma}_t$ follows

$$\mathbf{\Sigma}_t = \mathbf{H}\mathbf{P}_{t|t-1}\mathbf{H}^T + \mathbf{R}_t. \tag{S3}$$

Next, a prediction error, denoted $\tilde{\mathbf{y}}_{t|t-1}$, for the process model is calculated as

$$\tilde{\mathbf{y}}_{t|t-1} = \mathbf{z}_t - \mathbf{H}\hat{\mathbf{x}}_{t|t-1}, \tag{S4}$$

where $\mathbf{z}_t$ is a column vector containing the observed number of reported cases, hospital admissions, and reported deaths for day $t$ respectively in rows 1, 2, and 3. The data-updated estimate of the state variables, $\hat{\mathbf{x}}_{t|t}$ now follows as

$$\hat{\mathbf{x}}_{t|t} = \hat{\mathbf{x}}_{t|t-1} + \mathbf{K}_t\tilde{\mathbf{y}}_{t|t-1}. \tag{S5}$$

The data-updated covariance estimate, $\mathbf{P}_{t|t}$, satisfies

$$\mathbf{P}_{t|t} = (\mathbf{I} - \mathbf{K}_t\mathbf{H})\mathbf{P}_{t|t-1}, \tag{S6}$$

where $\mathbf{I}$ denotes the identity matrix. After setting the means and covariances of the variables $Z_r$, $A$, and $D_r$ in these updated estimates to zero, they can be used as initial values for projecting the process model forward to obtain $\hat{\mathbf{x}}_{t+1|t}$ and $\mathbf{P}_{t+1|t}$. In this way, process model predictions are obtained for all observations. Any missing observations, such as hospital admissions in data sets from early in the epidemic, are handled by setting the corresponding column in $\mathbf{K}_t$ to zero. Then, the results of these calculations may be used in an equation for the log likelihood.

The log likelihood of our model is calculated as the sum of the likelihood for the one-step ahead predictions of our process model and the step sizes in our random walk models for time-dependent parameters. The expression for the log likelihood of our process model predictions—*i.e.*, the marginal log likelihood of the Kalman filter—is

$$-0.5\sum_t [\tilde{\mathbf{y}}_{t|t-1,\text{nm}}^T \mathbf{\Sigma}_{t,\text{nm}}^{-1}\tilde{\mathbf{y}}_{t|t-1,\text{nm}} + \log\det\mathbf{\Sigma}_{t,\text{nm}} + d_{\text{nm}}\log(2\pi)], \tag{S7}$$

where the subscript 'nm', which stands for 'non missing', indicates that rows and columns which correspond to missing observations in $z_t$ have been omitted and $d_{\text{nm}}$ is the number of non-missing observations in $z_t$. The log likelihoods for the random walk steps are simply mean-zero Gaussian log-likelihoods with the appropriate variances.

### 2 Parameter initialization

In most cases, we initialize parameters by using theparameters estimated from a version of the observed data issued one week earlier for a particular location, which we refer to as a warm start. Any new random walk step parameters introduced by extension of the number of observations are assumed to be zero. In the first data set containing hospital admissions, the parameter $\tau_h$ is initialized with the sample variance of all non-missing observations, and the parameter $p_h$ is initialized as the quotient of the sum of all hospital admissions divided by the sum of all reported cases on days when hospital admissions were nonmissing.

In the first data set fitted, which in this study is the data set available on June 29, 2020, all parameter initializations had to be estimated from the data. We found that the following rough estimates lead to a satisfactory fit. All $\tau_{c,t}$ were intialized as the sample variance of cases divided by the mean number of cases. $\tau_d$ was initialized with the sample variance of observed deaths. If either of these values were less than 1, they were replaced with 1. An estimate of the unobserved time series of $Y$ was calculated by dividing the observed cases time series by $\gamma\rho_t \mathrm{d}t$, in which $\mathrm{d}t$ is the daily time step in our process model. The mean of the first 7 values of this time series times $\gamma/\eta$ was added to 1 to initialize $L_0$. The estimate of the time series of $Y$ times $\gamma p_h/\gamma_h$ was used to generate an estimate of the unobserved hospitalization time series. The parameter $p_d$ was initialized to the sum of observed deaths divided by the sum of these estimated hospitalizations. If this value was less than 0.01, the initialization was raised to 0.01. All $\gamma_{z,w}$ and $\gamma_{d,w}$ were initialized at the values given in Table 2.

An estimate of the time series of the effective reproduction number $\mathcal{R}_e = (\beta_t/\gamma)(X(t)/N)$ was calculated to generate initial values of $\beta_{\text{res}}$ and $\beta_{0,t}$. Let cases on day $t$ be denoted $c_t$. Our raw estimate of $\mathcal{R}_e$ was $(c_{t+7}/\rho_{t+7})/(c_t/\rho_t)$. These raw values were next clipped to be within 0.1 and 4 and then smoothed by applying a Savitzky-Golay filter of order 2 and length 21. To avoid bias from incomplete windows, the first and last 10 elements of output were considered missing. An estimate of a time series of $X$ was calculated by first initializing it with the same value that would be used in the process model given the initial value of $L_0$. Then, successive values were calculated by subtracting $c_{t+7}/\rho_{t+7}$ from the estimate of $X$ on day $t-1$. Then, a time series of $\beta_t$ estimates was calculated by multiplying the smoothed $\mathcal{R}_e$ estimate by $\gamma N$ and dividing by the estimate of $X$. Missing values on the ends of the time series were filled in by repetition of the first and last non-missing value. The slope from a linear regression of $\log \beta_t$ on $\texttt{residential}_t$ then provided the initial value of $\beta_{\text{res}}$. The residuals of this linear fit were grouped by week and averaged to provide the initialization of $\beta_{0,t}$. We have now described how all parameters were initialized.

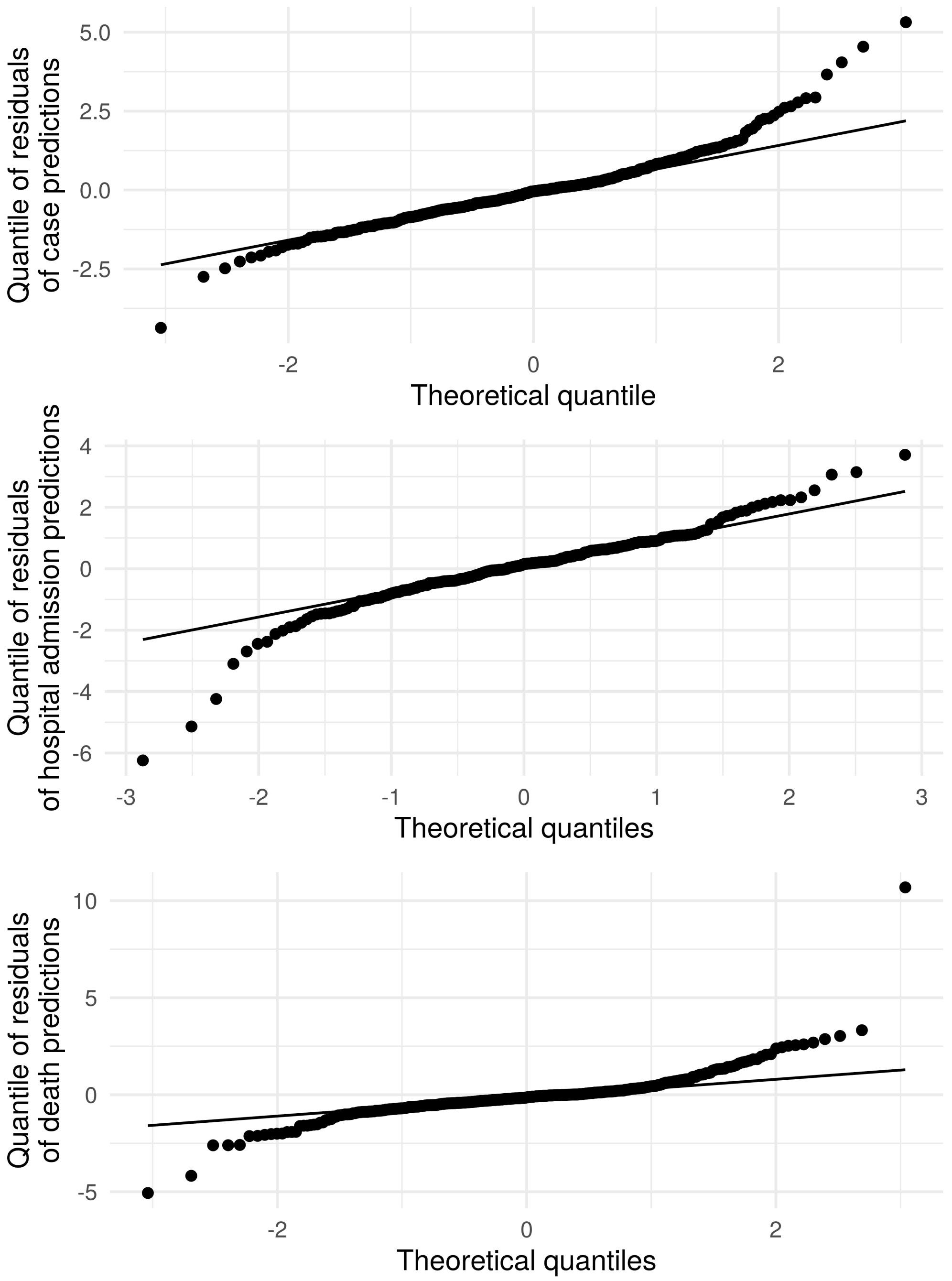


**Fig S1. Quantile-quantile plot of distribution of 1-step ahead forecast errors in fit to California data.** The specific residuals in the panels are the elements of $\tilde{\mathbf{y}}_{t|t-1}$ in equation S4 divided by the square root of variances on the diagonal of $\mathbf{\Sigma}_t$ in equation S3. These are quantiles from a fit to the data available on April 26, 2021, and thus represent a fit to data spanning the majority of the available data.

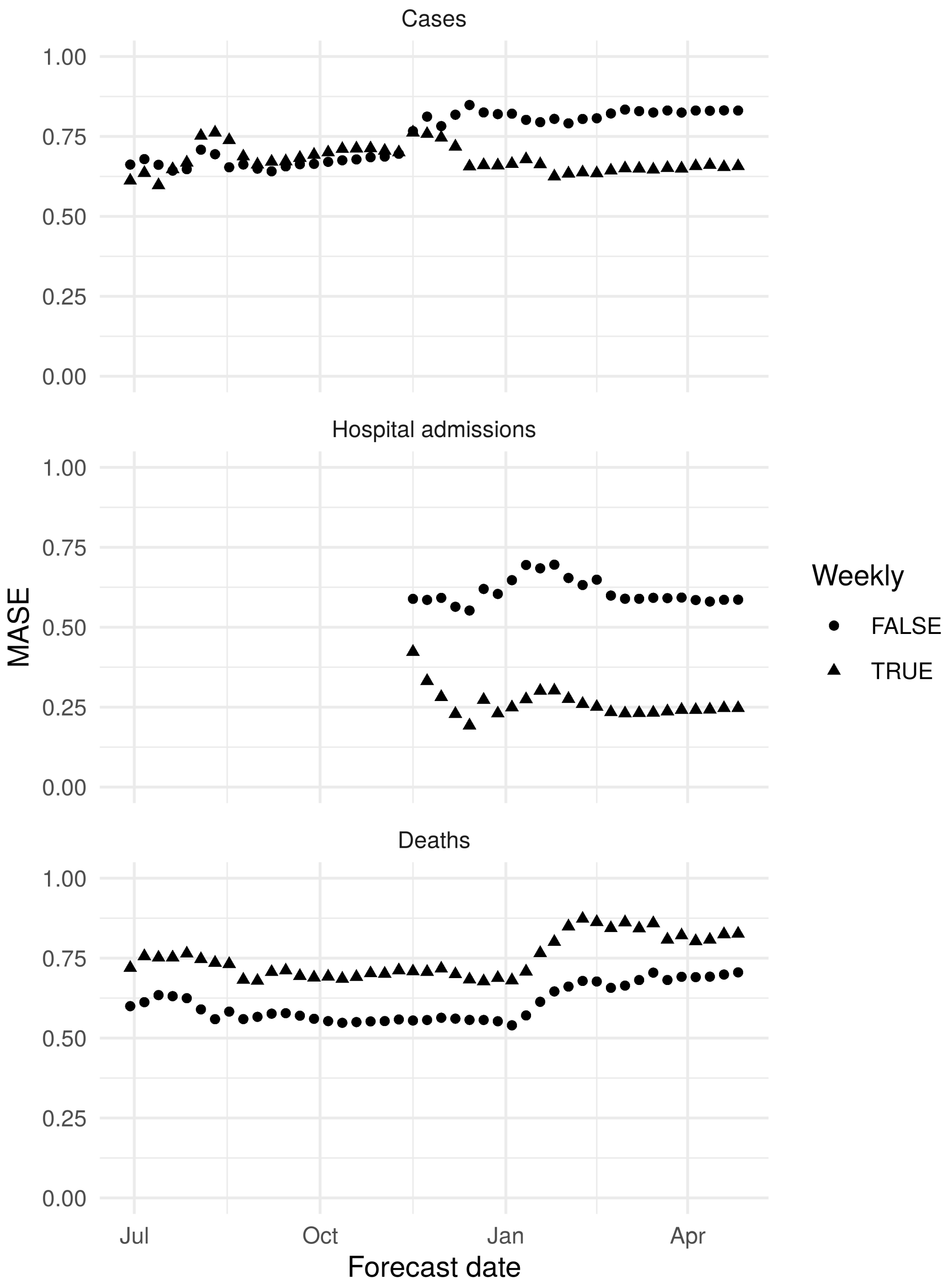


**Fig S2. MASE of fits to California data.** Weekly indicates whether MASE were calculated by using the previous observation from the same day of the week as the naive prediction (Weekly = TRUE) or whether MASE were calculated by using the previous day's observation as the naive prediction (Weekly = FALSE).

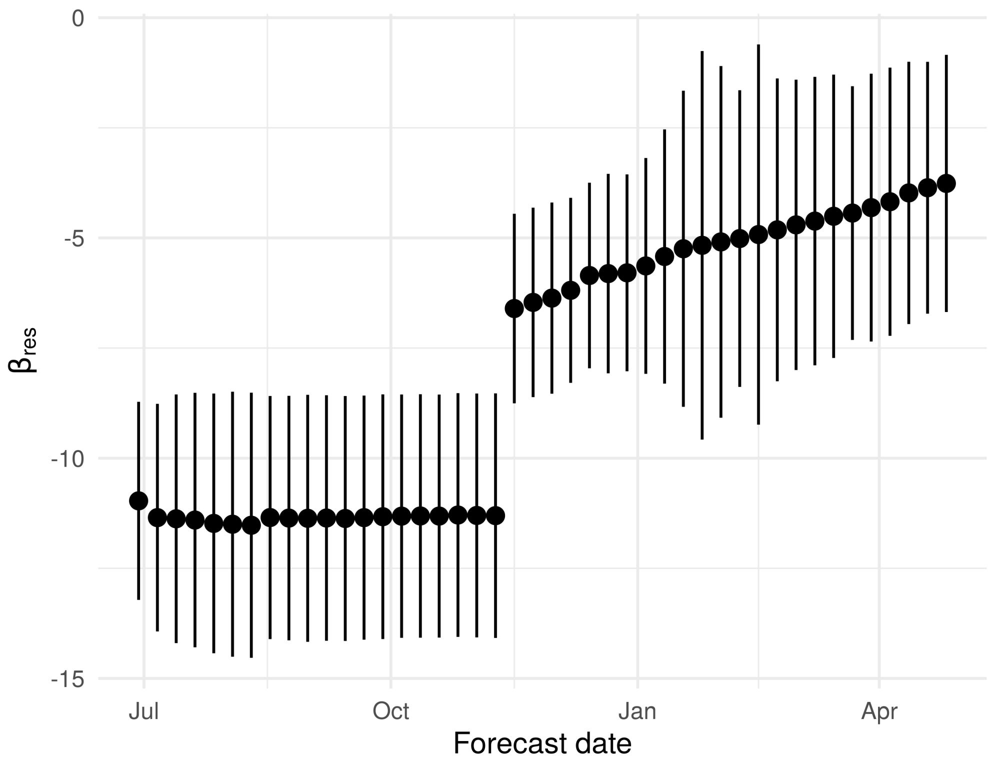


**Fig S3. The effect of time spent in residential areas on the transmission rate $\beta_{\mathbf{res}}$ changes as more data become available.** Error bars are 95% confidence intervals. Estimates come from modes fitted with data available up to the forecast date on the $x$ axis.

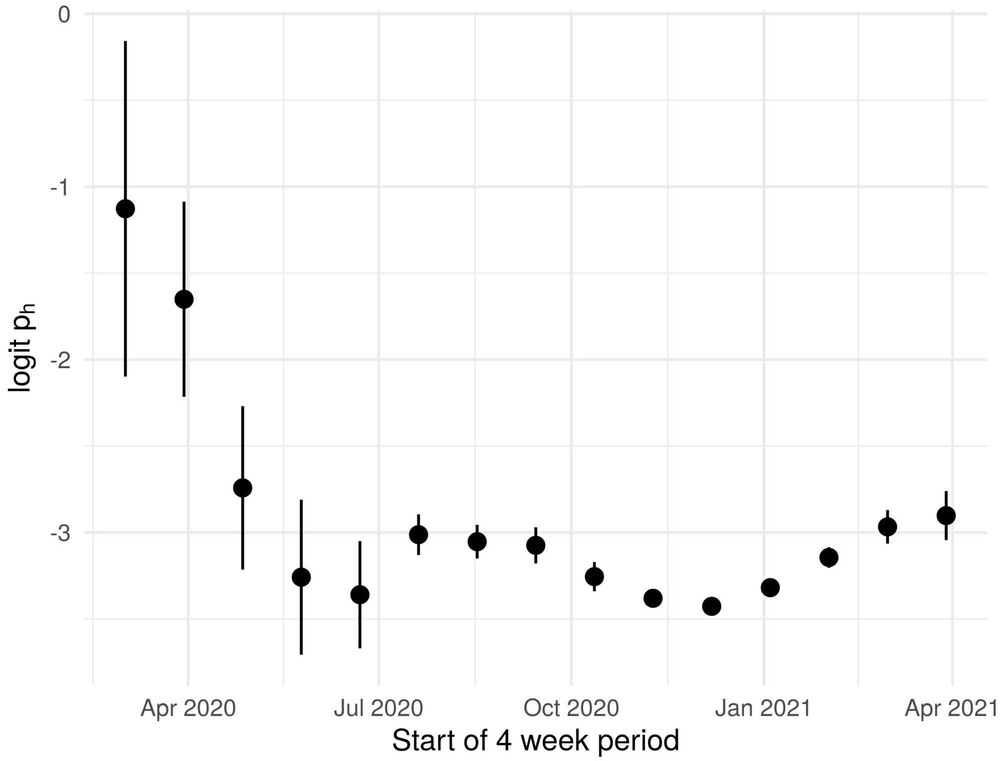


**Fig S4. Estimated probability of hospitalization $p_h$ over time in California model.** Points are maximum likelihood estimates and error bars are 95% confidence intervals.

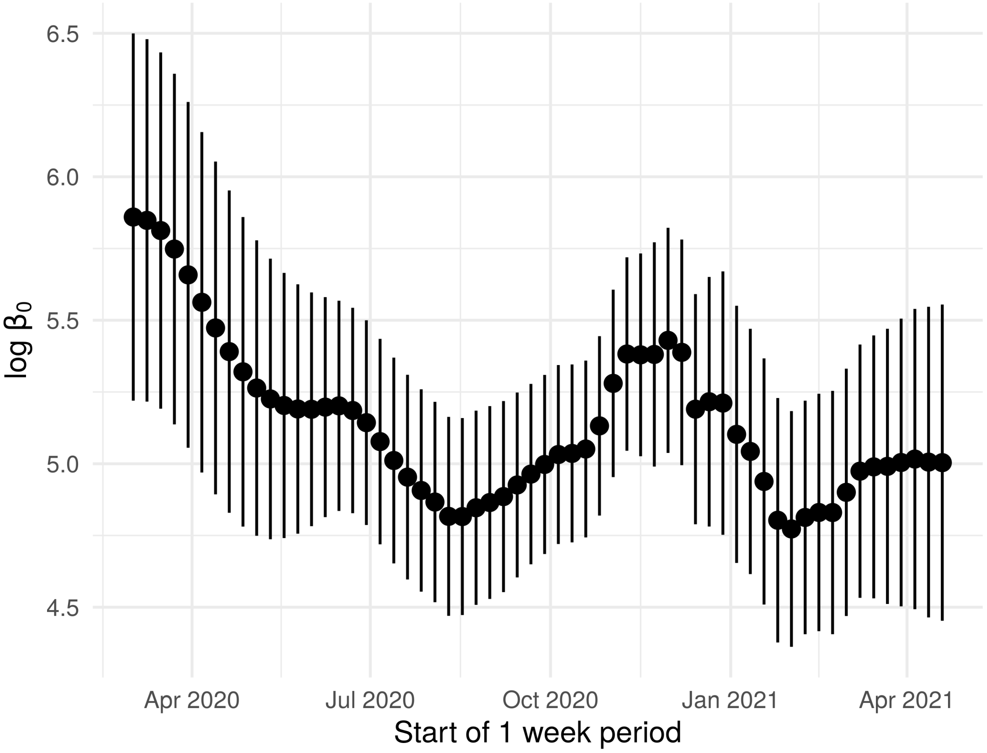


**Fig S5. Estimated transmission rate intercept $\beta_{0,t}$ over time in California model.** Points are maximum likelihood estimates and error bars are 95% confidence intervals.

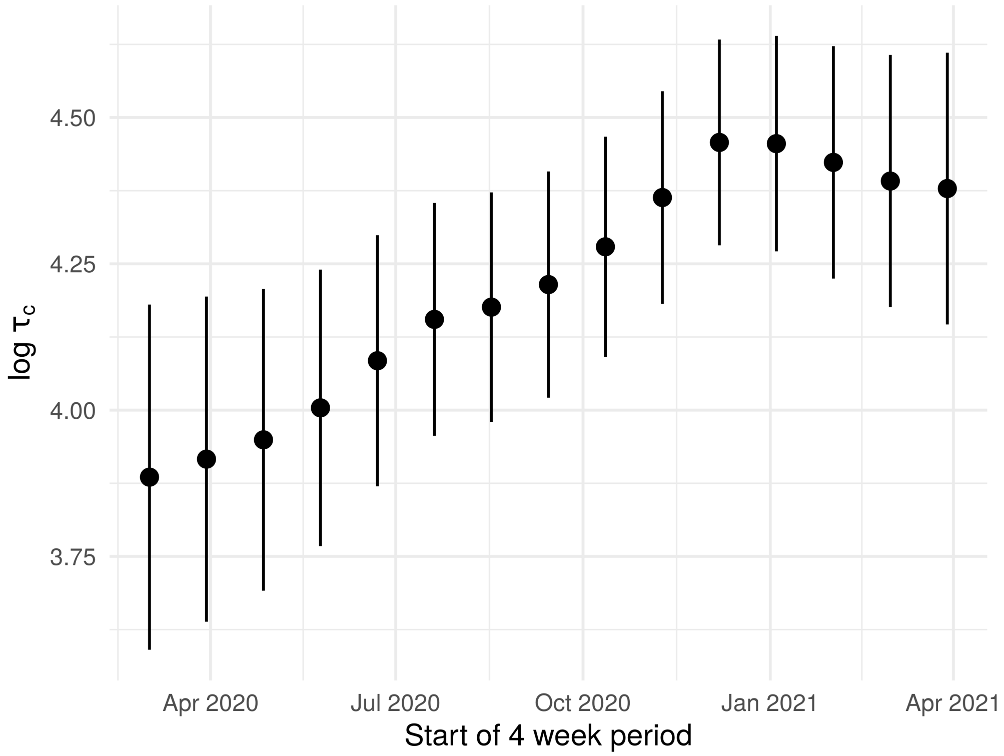


**Fig S6. Variance in observation error $\tau_c$ over time in California model.** Points are maximum likelihood estimates and error bars are 95% confidence intervals.

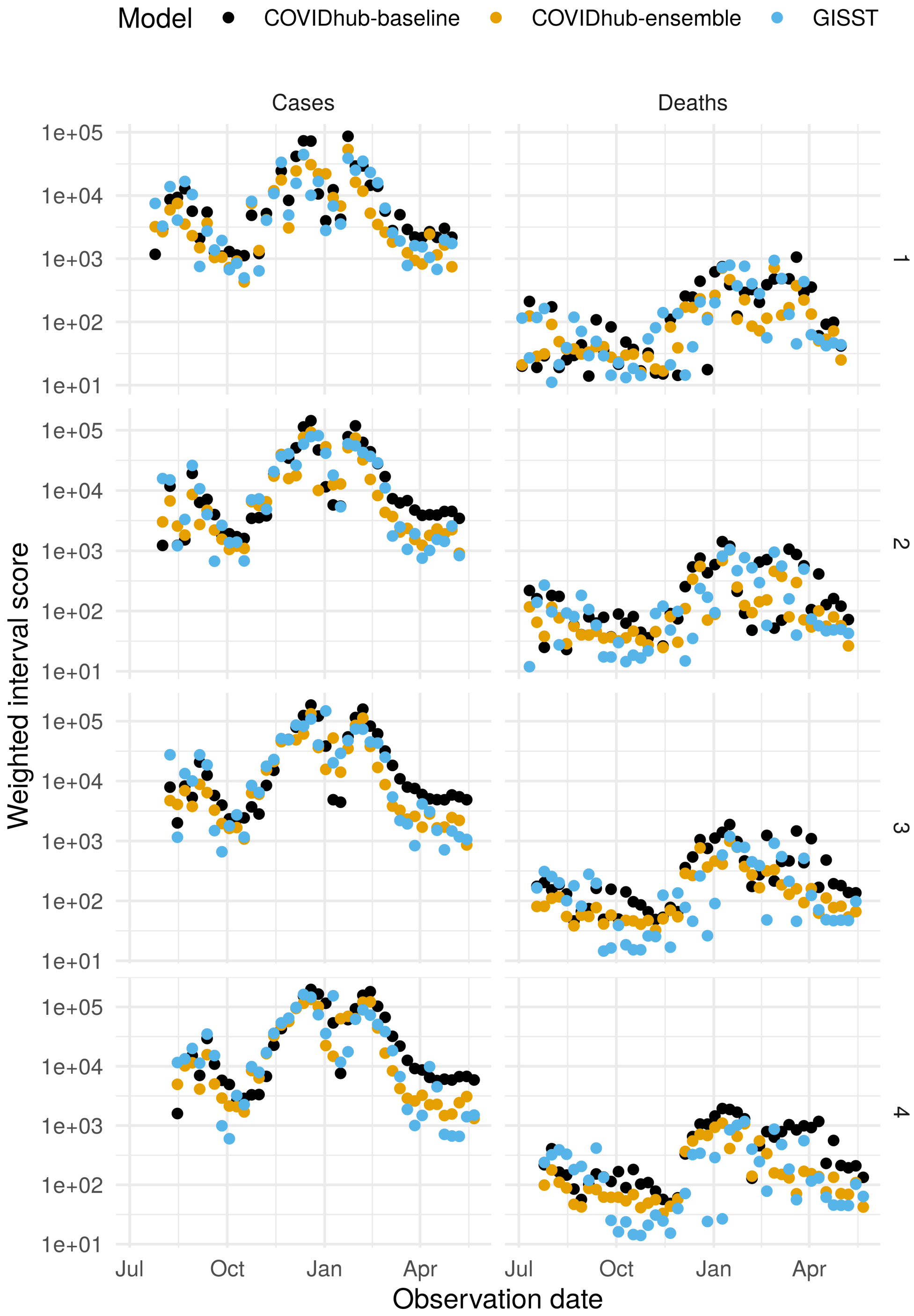


**Fig S7. Performance of short-term forecasts of cases and deaths by date.** The panel labels at right are the number of weeks in the forecast horizon. Lower scores indicate better performance.

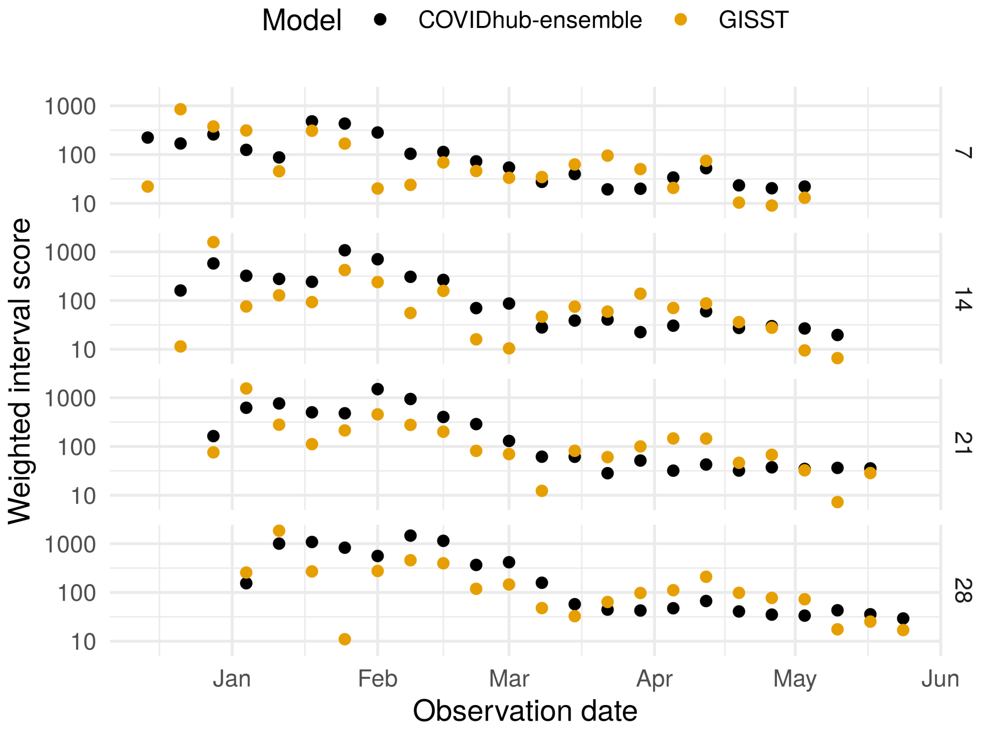


**Fig S8. Performance of short-term forecasts of hospital admissions by date for selected horizons.** The panel labels at right are the number of days in the forecast horizon. Lower scores indicate better performance.

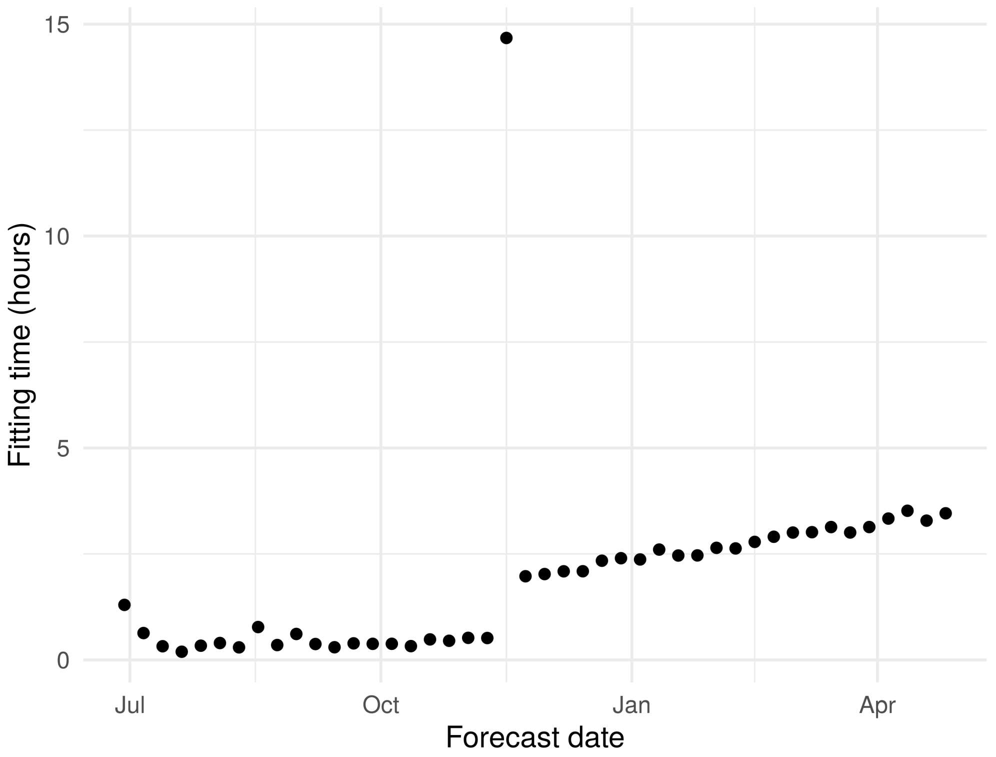


**Fig S9. Time required to estimate GISST parameters.** The spike in November was due to additional iterations due to the addition of hospital admissions data.
